# Building The Human Genotype-Phenotype Map to Harness Pleiotropy and Refine Disease Mechanisms

**DOI:** 10.64898/2026.02.19.26346618

**Authors:** Andrew R. Elmore, Aimee L. Hanson, Genevieve M. Leyden, James Johnson, George Davey Smith, Lavinia Paternoster, Tom R Gaunt, Gibran Hemani

**Affiliations:** NIHR Bristol Biomedical Research Centre, University Hospitals Bristol and Weston NHS Foundation Trust and University of Bristol, Bristol, UK; MRC Integrative Epidemiology Unit, Population Health Sciences, Bristol Medical School, University of Bristol, Bristol, UK

## Abstract

Mapping the pleiotropic effect of genetic variation on biological processes and complex phenotypes is fundamental to extracting translational insight from genome-wide association studies (GWAS). Here we present The Human Genotype-Phenotype Map (GPMap), a repository of colocalizing genetic associations across 15,997 complex traits and 2.7 million molecular measurements, leveraging common and rare variants and *cis*-and *trans*-acting effects across disaggregated tissue types and single cell datasets to trace the complex pathways through which they act. We identify over 49.3 million colocalizing trait pairs, which aggregate into 97,393 colocalization groups, representing distinct pleiotropic variants based on shared genetic signals, with 55.8% of genome-wide significant disease-associated loci colocalizing with at least one molecular trait. This insight facilitates clustering of complex health and disease phenotypes based on genetic architecture, and the dissection of polygenic traits reflecting the composite impact of many underlying processes. We show that leveraging pleiotropic information can enhance the selection of genetic instruments for causal inference approaches and improves prediction of drug trial success. This open-source resource is available at https://gpmap.opengwas.io, with functionality for user GWAS upload.

## Introduction

Uncovering molecular mechanisms and shared biology for complex traits underpins the motivation for the past two decades of genome-wide association studies (GWAS). Much of the insight and interpretation needed to achieve these goals is gained by leveraging the phenomenon of pleiotropy, in which a single genetic variant can influence multiple traits (molecular and phenotypic). Mapping pleiotropy is a fundamental component of building genotype-phenotype maps, aiming to capture the mechanistic translation of genetic polymorphisms to phenotypic consequences. The degree to which pleiotropy manifests universally, whereby each variant potentially influences all traits, or modularly, by which variants acting within a given biological pathway predominantly modulate a specific set of traits, has been a point of discussion with implications for species evolvability for over a century(1–5).

Pleiotropy has been defined in a multitude of ways depending on the mechanistic context under investigation (**Supplementary note 1**). Variant-level pleiotropy mapping aiming to determine shared causal variants is routinely performed in genetic epidemiology studies to gain greater insight into trait mechanism, typically examining hypothesis-driven trait relationships. For example, genome-wide sharing of genetic effects between traits is routinely estimated to infer overlapping and non-overlapping aetiological mechanisms, and variant function is often revealed through multi-trait relationships discovered through phenome-wide association studies.

Pleiotropy mapping has been used to boost statistical power in GWAS by leveraging shared genetic signals between related traits(6). In addition, causal genes for traits are often detected through colocalization analysis(7), whereby assessment of the linkage-disequilibrium (LD) structure at loci shared between molecular and complex traits of interest aids differentiation of proximal associations from those reflecting the same causal signal. Such an approach is frequently used in drug target prioritisation efforts(7). Modelling pleiotropy is also fundamental to the Mendelian randomization (MR) analytical framework for examining causal relationships between traits. The essence of estimating causal relationships in MR is in modelling vertical pleiotropy, defined as the genetically proxied putative causal factor having an effect on the outcome proportional to its effects on the exposure(8). These causal estimates are biased should variants influence the outcome through pathways that exclude the exposure (i.e. through horizontal pleiotropy). Numerous methods have been developed to manage such biases, increasingly through detecting pleiotropic mechanisms and adjusting for them directly (9,10).

Inherent constraints in data harmonisation, computational scalability, and modeling complexity frequently limit the depth of pleiotropy mapping across these different study designs. One major challenge is in collating all relevant traits for the study, and another is in appropriately modelling the sharing of loci across those traits. While the human phenotypic space is essentially boundless in terms of the traits that can be defined and the contextual dimensions in which they can be measured, the breadth of measured molecular and phenotypic traits with publicly available GWAS summary statistics from well-powered biobanks has grown dramatically and with an accelerating trajectory. Here we describe efforts towards a putative genotype-phenotype map (GPMap) that systematically maps pleiotropy among 15,997 complex traits, including physiological measures and disease endpoints, and 2.7 million molecular measurements from tissue specific and single cell datasets. We have performed common variant summary statistic imputation, fine-mapping and colocalization for all extracted traits, and utilized graph-based methods to group traits with a high probability of sharing a causal signal (**Figure 1**). Where available, we also report on shared rare variant associations. We demonstrate the power of the GPMap in capturing the breadth of pleiotropy underlying human phenotypes, supporting mechanistic insights into drivers of health and disease for drug target prioritisation, and enhancing instrumental variable (IV) selection for causal inference. The resource is continually updated with the availability of new GWAS summary data, and is made accessible through a web application, R package and direct download.

**Figure 1:**
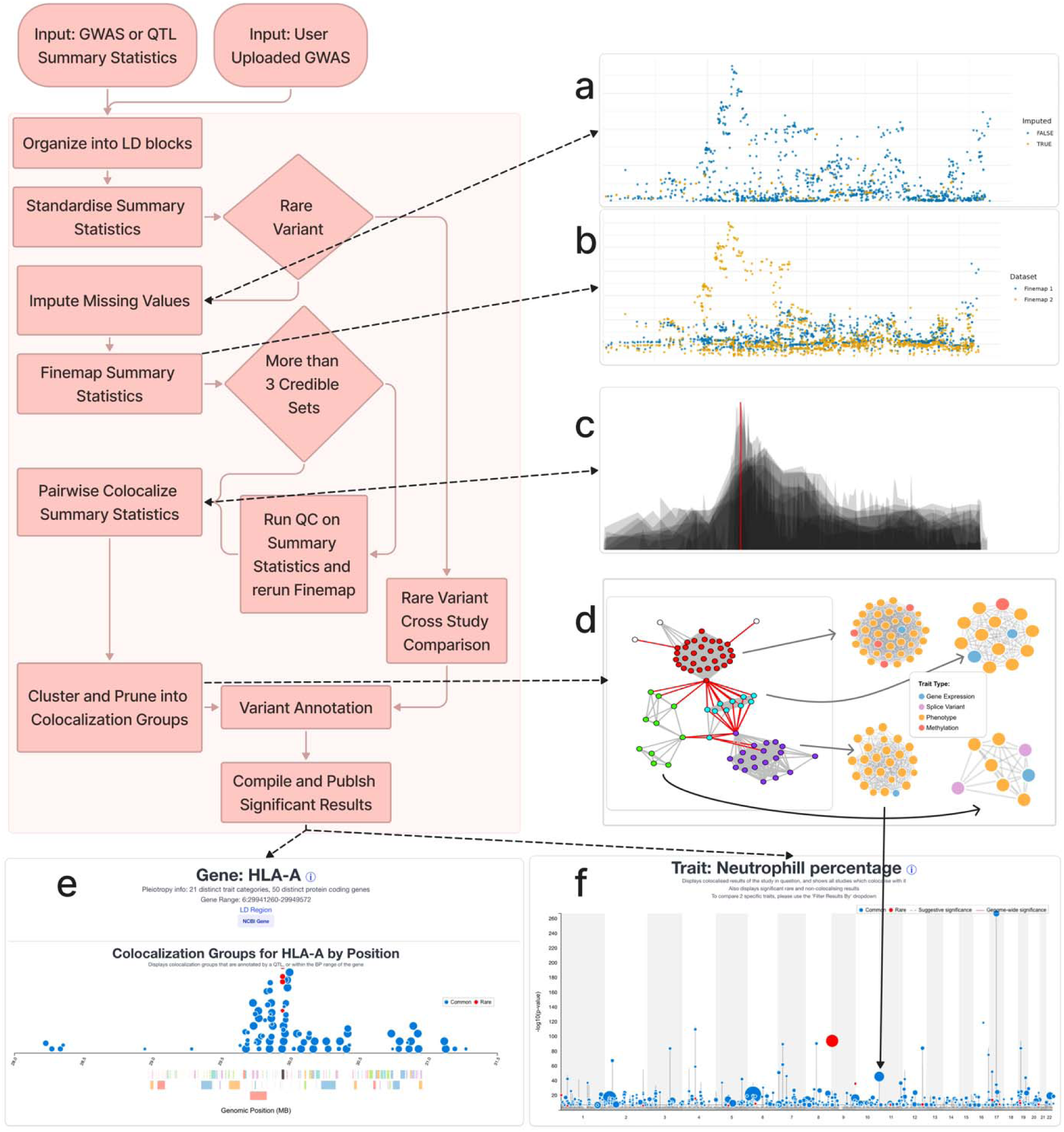
The Genotype-Phenotype Processing Pipeline. The input of the pipeline is a set of GWAS summary statistics, a series of steps is then performed to standardize the data and isolate the causal variants, which are then compared via colocalization and clustered into groups. These are then available to be investigated via a series of views: via trait, gene, variant, or tissue. The images are an illustration of the pipeline and is not representative of real data in the GP Map. a. GWAS summary statistic imputation step: where missing variants from the reference panel are imputed from data from the LD reference panel b. Fine-mapping Step: where we ran fine-mapping on each LD block where there exists a variant where p<1.5e-4. The log bayes factor was stored. c. Colocalization step: where the log bayes factor data was used to run colocalization against any two traits d. Clustering and pruning step: where a graph of all traits as node and significant colocalization results as edges is created, and a graph-base clustering and pruning algorithm found colocalization groups. In this example, edges marked in red were classified as false positives, and the loosely connected cluster was split into 4 colocalization groups. e. Result amalgamation (by gene): where colocalization and rare variant results were amalgamated by all molecular traits that were annotated by gene. f. Result amalgamation (by trait): where colocalization and rare variant results were amalgamated by trait.

## Results

### Data and Results Overview

The GPMap processing pipeline (**Figure 1**) was developed to extract publicly available GWAS summary statistics from 12 sources (**Figure 2a**, **Supplementary Table 1**) and systematically maps shared genetic associations across all traits at 5.26 million distinct loci with a suggestive significance threshold (p<1.5e-4). For common variants, summary statistic imputation was performed on standardized input data, followed by fine-mapping and pairwise trait colocalization. Traits with colocalizing genetic signals at each locus (H_4_ probability of a shared causal variant ≥0.8) were grouped into multi-trait clusters, which enabled inference of false positive and false negative colocalization results and further cluster segregation. Shared rare variant signals were also reported for a subset of traits. For further explanation of the processing pipeline, see **Methods**, for term glossary and definitions, see **Supplementary Note 1**. The GPMap further supports input of user provided GWAS summary statistics, enabling custom integration into the broader pleiotropy landscape.

**Figure 2:**
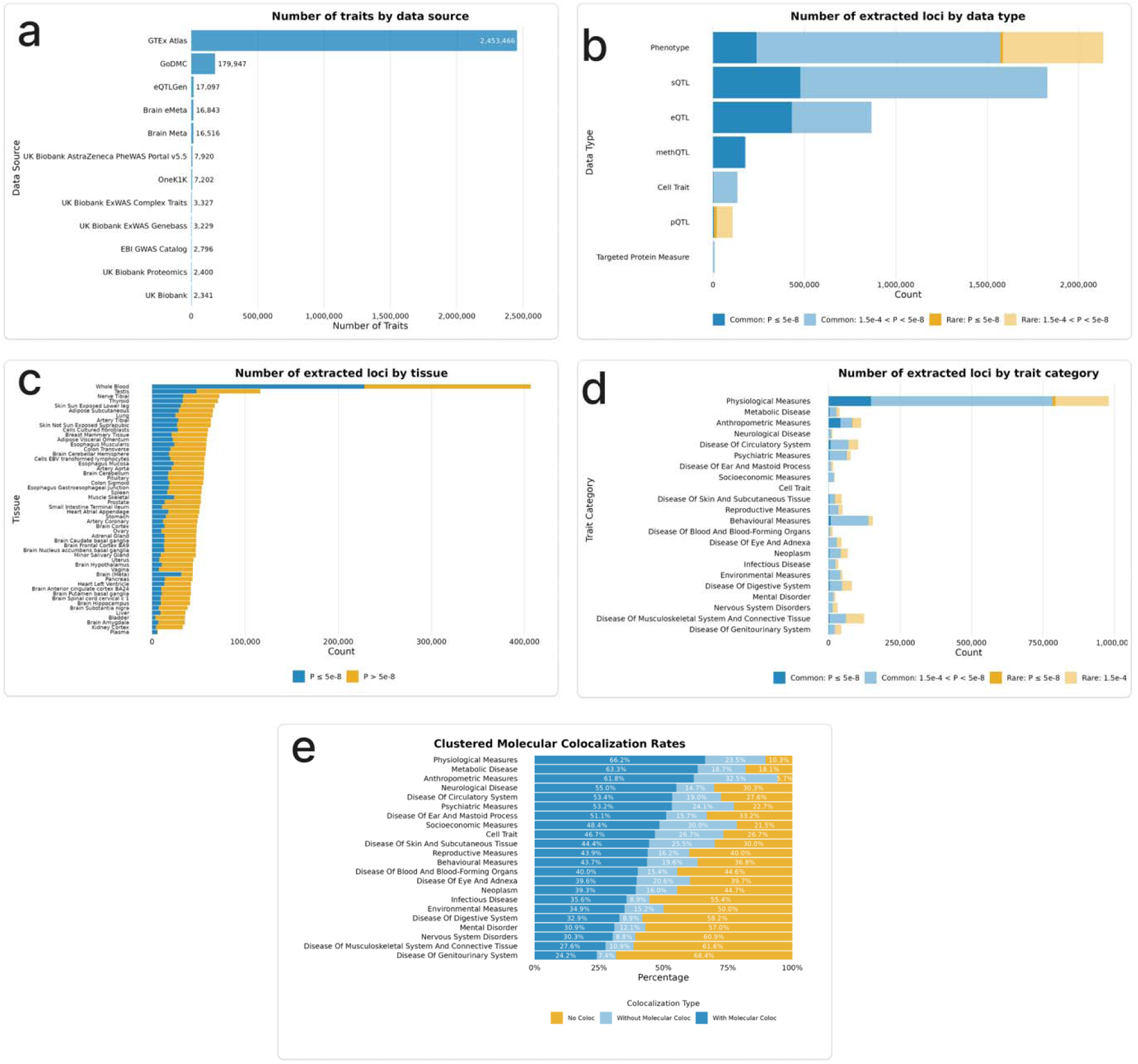
Summary of Data Each graph presents a distinct aggregation of the input data and its associated colocalization outcomes. a. A summary of the number of traits by data source. b. A summary of the complex and molecular loci extracted by data type. c. A summary of the number of extracted loci by tissue, split by genome-wide significant p-value threshold, and by variant frequency (where a variant < 0.1% is considered a rare variant). d. Number of extracted loci by trait category. A further breakdown of the ‘phenotype’ category. e. Clustered Molecular Colocalization Rates: The molecular colocalization rates of each significant colocalization result (H4≥0.8) after clustering, broken down by trait category. The clustering marked some results as false positives and false negatives. The rates of molecular colocalization slightly increased across all trait categories. QTL: Quantitative Trait Locus

All contributing loci as suggestive significance (p<1.5e-4) included 2.13 million associations across all complex traits (1.57 million common variants from 4,475 traits at minor allele frequency (MAF) >1% and 563,567 rare variants from 11,522 traits at MAF < 1%), and 3.13 million molecular quantitative trait loci (QTL) from studies assaying mRNA exon splicing (sQTL), mRNA expression (eQTL), CpG site DNA methylation (methQTL) and plasma protein levels (pQTLs; 2.01 million common variants and 102,126 rare variants, **Figure 2b**). There were 1.36 million loci reported as genome-wide significant (GWS; p<5e-8) in at least one trait. The integrated QTLs enabled loci to be annotated to 38,372 genes and pseudogenes across 53 tissue types and 14 cell types (**Figure 2c**), and complex traits were organized into 23 trait categories spanning disease subclasses and physiological measures (**Figure 2d**).

Collectively these per-trait loci aggregate into 97,393 independent groups of colocalizing traits, referred to herein as ‘colocalization groups’ (**Figure 1d**), refined from 49.3 million individual pairwise trait colocalization results (H_4_≥0.8). Further, we identified 1,053 GWS, and 217,636 p<1.5e-4 rare variants shared by more than one trait, referred to herein as ‘rare variant groups’. We found colocalization methods to be susceptible to making false positive and false negative H_4_ calls, particularly in regions of long-range LD, sparse variant coverage or low study power.

Likely erroneous trait connections were only identifiable upon linking traits into multi-trait graphs, enabling pruning of sparse links between dense trait clusters and inference of missing colocalization calls (**Supplementary Note 2-3**). The graph-based clustering process demonstrated high internal consistency, with an average group connectedness of 85.5%. 4.0% of all pairwise colocalizations with H_4_≥0.8 at GWS were classified as false positives, as they lacked sufficient connectivity within their prospective colocalization group. Conversely, 14.5% of GWS traits with H_4_<0.8 were identified as false negatives that were subsequently recovered through their inclusion in high-confidence clusters.

Overall, 81.4% of the 276,804 GWS common complex trait loci analyzed colocalized with at least one other trait. 55.8% of GWS complex trait loci colocalized with at least one molecular trait locus (though this varied by complex trait category, **Figure 2e**). This colocalization rate marks a notable increase over previous estimates of 43% (11), underscoring the power of our resource yielded by extensive phenome coverage, and the recovery of false negative colocalization calls through simultaneously leveraging millions of trait-trait relationships. For further insight into different types of CGs, see **Supplementary Note 4**.

### Colocalization Distinguishes Causal Variants from Confounding due to LD

While shared GWAS signals indicate that two traits are associated with the same genetic region, colocalization provides statistical evidence to determine if they are driven by the same underlying causal variant. Our results demonstrate a marked disparity between proximal signal sharing and colocalization, further supporting that colocalization is a necessary prerequisite in establishing a causal relationship.

We evaluated the prevalence of biological versus LD-induced pleiotropy by comparing the ratio of trait regions that had evidence for a signal (p<1.5e-4) within a ±1Kb window of the causal variant as calculated by SuSiE(12), but did not share a causal variant (H_3_≥0.8) to those that that did share a causal variant (H_4_≥0.8) was 0.29:1. This indicates that ∼22% of colocalizing associations prioritized by proximity alone represent distinct genetic architectures rather than true pleiotropy. Such a high prevalence of spurious overlaps suggests that a substantial fraction of instrumental variables (IVs) may lead to erroneous functional interpretations if not rigorously filtered through colocalization frameworks(13).

### Characterization of Pleiotropy and LD

Analysis of the mapped pleiotropy landscape corroborated a previously hypothesized L-shaped distribution at the variant level (14–16), characterized by the majority of loci exhibiting specific effects on a trait category, and a subset of variants demonstrating associations across multiple trait domains. Notably, we are able to examine this pattern in different dimensions. We find this L-shaped distribution in the size of colocalization groups, with most having few traits (**Figure 3a**), the number of trait categories associated with a specific gene (**Figure 3b**), as well as the number of protein coding genes (annotated through QTL signals) within a given colocalization group (**Figure 3c**).

**Figure 3:**
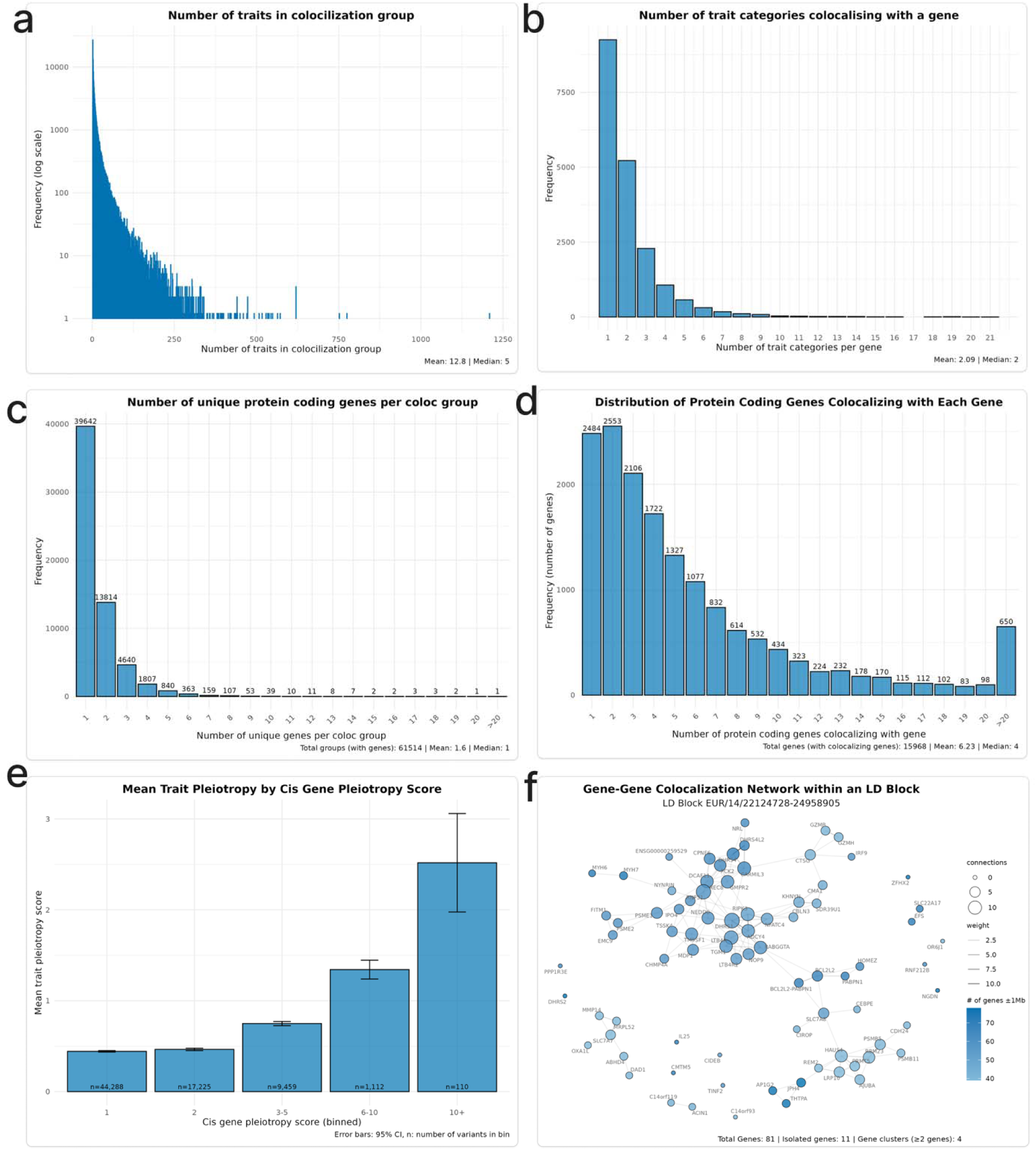
Investigation into Pleiotropy Using the Genotype-Phenotype Map Each graph investigates a different lens into the L-shaped distribution of pleiotropy, where many genes are oligopleiotropic, meaning they affect only one or a few traits (the tall part of the’L’), few genes are highly pleiotropic, meaning they affect a large number of traits (the short, long tail of the’L’). a. Histogram of the frequency of colocalization group size. b. Number of genes colocalizing per trait: This show the inverse relationship to (A), where most traits are associated with a small number of genes, and some traits are highly pleiotropic. c. Number of Trait Categories Colocalizing with a Gene: For each gene, the number of trait categories that gene colocalizes with is summed. The frequency is how often any given gene has the number of trait categories on the y-axis. Most genes are associated with a single trait category (oligopleiotropic), and some are associated with many (highly pleiotropic). d. Number of Protein Coding Genes per Colocalization Group: For each colocalization group, the number of unique protein coding genes are summed. The frequency is how often any given colocalization group has that number of genes in it. e. The mean number of distinct trait categories for variants binned by their cis gene pleiotropy score, revealing that variants tagging a greater number of cis genes tend to be associated with a broader range of trait categories. f. Gene-Gene Colocalization Network within an LD Block: Each colocalization link between one gene and another is summed and displayed as a network graph. This shows a modular relationship between proximal genes within a single LD block.

Mendelian randomization using gene-level molecular traits as exposures (such as transcript expression or protein levels) tend to assume that *cis-*QTLs (falling within a given base pair window around the instrumented gene) are specific to the gene. However, we demonstrate that most genes share *cis-*QTLs with at least one other gene in the *cis* window of that gene (**Figure 3d**). Though colocalization probability between genes increased with physical proximity, genomic distance was a poor predictor of colocalization events between genes (β=0.072, R^2^=0.161, p=0). While colocalization mitigates linkage-driven artifacts, we observed that the detection of trait category pleiotropy scales with the number of independent molecular traits that are measured for a gene (**Figure 3e**). This suggests that variant-level pleiotropy may be driven by a variant influencing independent pathways through perturbing multiple genes in a region, more than the phenomenon of single genes impacting multiple trait domains.

Finally, we examined the gene-gene colocalization network of a representative genomic region (chr 14: 22124728-24958905, **Figure 3f**), revealing that colocalizing molecular signals can link many genes in pleiotropic networks, as well as identify variants influencing single genes in isolation. As demonstrated below, leveraging gene-gene and gene-trait networks constructed in this way enables the following: assessment of the biological specificity of genetic variants, mapping of pathways linking genes to complex traits, and prediction of the potential consequences of therapeutics interfering with such pathways.

### The GPMap uncovers the genetic architecture of hemoglobin levels

To demonstrate the value of integrating phenome-wide GWAS evidence into a GPMap, we investigated the heterogenous pathways influencing hemoglobin concentration through genetic pleiotropy. Anchoring on a GWAS of this measure in UK Biobank (https://gpmap.opengwas.io/trait.html?id=4759; hereafter taken as the “anchor trait”), colocalizations were observed at 447 independent variants (linked to 773 genes), capturing shared genetic architecture across 6734 traits (983 complex traits and 5751 molecular traits; **Figure 4a, Supplementary Table 2**). A further 22 rare variant associations (linked to 304 genes) were shared between the anchor and 653 traits (334 complex traits, 318 protein QTLs (pQTL); **Figure 4a, Supplementary Table 3**). As expected, the common variant traits showing the greatest proportion of colocalizing GWS associations were for highly correlated or related measures (e.g. hematocrit, red blood cell count and liver iron content), but multiple genome-wide colocalizations were also observed for complex traits expected to be more peripheral causes (e.g. autoimmune disease(17)) or consequences (e.g. low birth weight(18)) of altered hemoglobin levels (**Figure 4b**, **Supplementary Table 4-5**).

**Figure 4:**
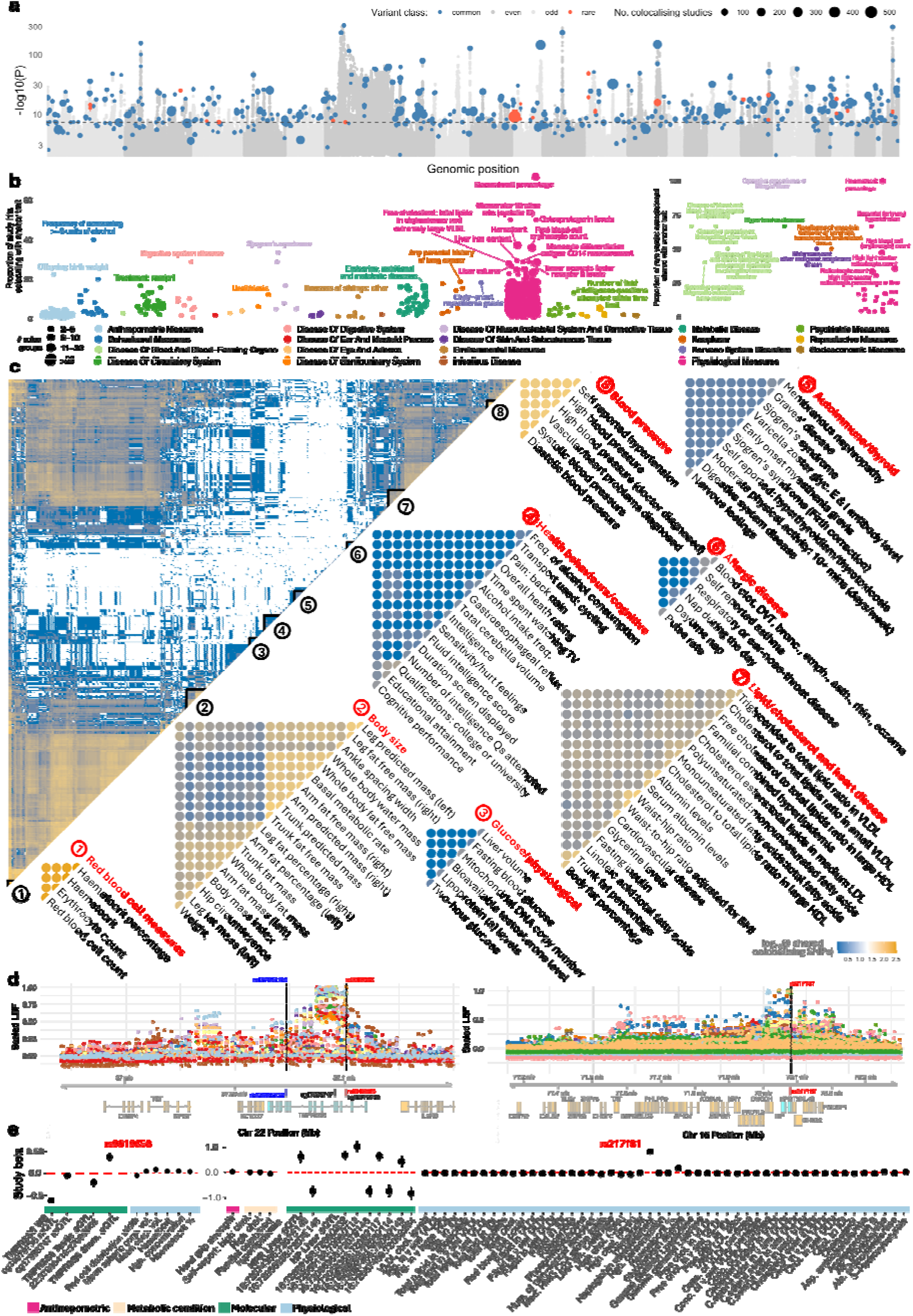
Mapping the contributors to blood hemoglobin concentration. a. Manhattan plot of association summary statistics from a UK Biobank GWAS of blood hemoglobin concentration (the anchor GWAS; OpenGWAS: ukb-d-30020_irnt). Fine-mapped common variants which colocalize with other GPMap complex traits are colored blue, with points scaled by the number of studies which share the colocalizing variant. Only colocalizing studies with least one fine-mapped signal passing genome-wide significance (GWS) in the colocalized region were retained, although the chosen candidate variant representing the study group may not itself exceed GWS in all studies. Rare variants shared between the anchor trait are other phenotypes are superimposed on the plot and colored red. Even and odd chromosomes 1-22 are shown in alternating grey. b. The proportion of all independent GWS study hits which colocalize with the anchor GWAS, restricting to studies of complex traits which show at least 2 independent colocalizations with the anchor. Points are scaled according to the number of colocalizing hits (i.e. the number of colocalization groups both studies occupy), and colored according to broad trait categories. An equivalent plot for shared rare variants is shown to the right. c. Hierarchically clustered heatmap of the number of shared colocalizing hits between any two pairs of studies and the anchor trait. Example groups of studies which share common genetic signals are shown and assigned broad trait categories. Low sharing of signals between groups indicates heterogeneity in the genetic factors that contribute to hemoglobin levels. d. Example of colocalizing complex and molecular trait signals at two hemoglobin associated loci, *TMPRSS6* (left) and *HP/HPR* (right). Locus plots show log bayes factors (LBF) returned from SuSiE fine-mapping, scaled by the maximum LBF observed across all studies in the colocalization group. Colored points represent the different studies colocalizing at each locus, with the position of the assigned candidate variant (the variant with the highest cumulative LBF across studies) annotated in red and shown relative to genes in the locus below. For the TMPRSS6 locus, the location of two CpG sites that associate with the candidate SNP are shown, along with a rare variant previously associated with severe iron refractory iron deficiency anaemia, annotated in blue. e. Effect estimates for the candidate SNPs shown in d. for studies in the respective colocalization groups. All associations pass GWS. Error bars represent 95% confidence intervals. Duplicate and highly correlated traits have been removed for visualization purposes; all betas are presented in **Supplementary Table 7.** Ala=alanine, alk=alkaline, asp=aspartate, chol.=cholesterol, conc.=concentration, stom.=stomach, hgb.=hemoglobin, p.lipids=phospholipids, p.glycerides=phosphoglycerides, t.aminase=transaminase, t.peptidase =transpeptidase, trig=triglycerides, VLDL=very low-density lipoprotein, LDL=low-density lipoprotein, HLD=high-density lipoprotein, IDL=intermediate-density lipoprotein.

We performed hierarchical clustering of the pairwise complex trait matrix enumerating the number of colocalizing variants between each complex trait pair and the anchor trait, which identified distinct groups of traits with similar colocalization patterns.

Physiological/anthropometric traits, such as erythrocyte measures, lipid and cholesterol levels, body size parameters and blood pressure, showed strong clustering into distinct groups (**Figure 4c**), which showed little genetic overlap with independent clusters of colocalizing traits related to cognition and health behaviours, autoimmunity and allergic disease. Through this approach, multi-trait colocalization and clustering can help disentangle the complex amalgamation of genetic signals underlying a multifaceted complex trait.

We next investigated genetic signals from two colocalization groups containing the anchor trait alongside molecular QTLs for genes related to iron homeostasis and hemoglobin recycling. The first colocalization group, represented by the candidate variant chr22:37100807 C/T (rs9619658; **Figure 4d**), an intronic variant in *TMPRSS6,* associated with decreased methylation at two CpG sites within *TMPRSS6* (cg17956747, 22:37095554-37095556; cg19979738, chr22:37100888-37100890), alternate expression of a *TMPRSS6* splice variant and increased *TMPRSS6* gene expression, as well as increased red blood cell volume/hematocrit and blood hemoglobin concentration (**Figure 4e** left; **Supplementary Table 6**). *TMPRSS6* is a transmembrane serine protease and important negative regulator of the primary iron regulatory hormone hepcidin(19). We identified a shared pathogenic rare variant in *TMPRSS6,* which has been associated with increased hepcidin expression, reduced uptake of dietary iron, low hemoglobin and severe iron-refractory iron deficiency anaemia(20,21) (shown in blue in Figure 4d). Common polymorphisms that increase *TMPRSS6* activity act as genetic modifiers to amplify iron overload in hereditary hemochromatosis(22).

The second example captures colocalizing signals represented by candidate variant chr16:72080103 C/T (rs217181), situated downstream of the haptoglobin (*HP*) and haptoglobin receptor (*HPR*) genes. Here we see colocalization between *HP* and *HPR* splice variants detected in liver and blood, increased plasma haptoglobin levels, reductions in cholesterol, phospholipid and triglyceride levels in low and very low-density lipoproteins (LDL; VLDL), low hemoglobin concentration and protection from high cholesterol and hyperlipidemia (**Figure 4d and 4e** right; **Supplementary Table 7**). These observations are consistent with the known, duel independent roles of HP; binding hemoglobin to prevent toxicity and iron loss during red blood cell turnover(23), and binding apolipoprotein E (ApoE) to prevent ApoE oxidation, aiding ApoE-mediated plasma lipid clearance and reducing total and LDL cholesterol levels(24). Copy number variation in *HP* C-terminal exons, tagged here by GTEx(11) liver sQTLs, differentiate two protein isoforms (HP1 and HP2) which differ in their ApoE antioxidant capacity and strongly associate with cholesterol levels(25,26). Thus, assessing the colocalization of extensive molecular, physiological and health related phenotypes at specific genomic sites can reveal layers of genomic regulation, including complex alternate isoform usage, underlying the pleiotropic role of genes involved in hemoglobin regulation as well as disparate biological processes.

### Tissue-annotated colocalization signals facilitate phenotypic refinement

Complex trait heterogeneity often stems from traits representing composite biological endpoints. Integrating tissue-specific gene expression can help refine these mechanisms, improving both our understanding of trait etiology and selection of instrumental variables (IVs) to proxy exposures for MR (27–29). We show how the GPMap may be leveraged to refine biologically distinct sets of genetic variants stratified by tissue-type using body-mass index (BMI, https://gpmap.opengwas.io/trait.html?id=1992) as an example endpoint trait.

BMI is a composite complex trait capturing distinct biological processes involving different body systems. We hypothesised that the traits colocalizing at BMI loci would therefore differ depending on the tissue-specificity of the QTL signals observed (its tissue annotation). 992 candidate variants represent colocalization groups that include BMI, of which 320 have evidence for colocalization with tissue-derived eQTL. **Figure 6a** displays the top 18 tissues represented in BMI colocalization groups. This highlights that BMI shares the most loci with brain-tissue derived eQTL (115 loci), of which 41 are specific to brain-tissue, while a large proportion of BMI loci colocalize with eQTL not specific to any one tissue type.

**Figure 5:**
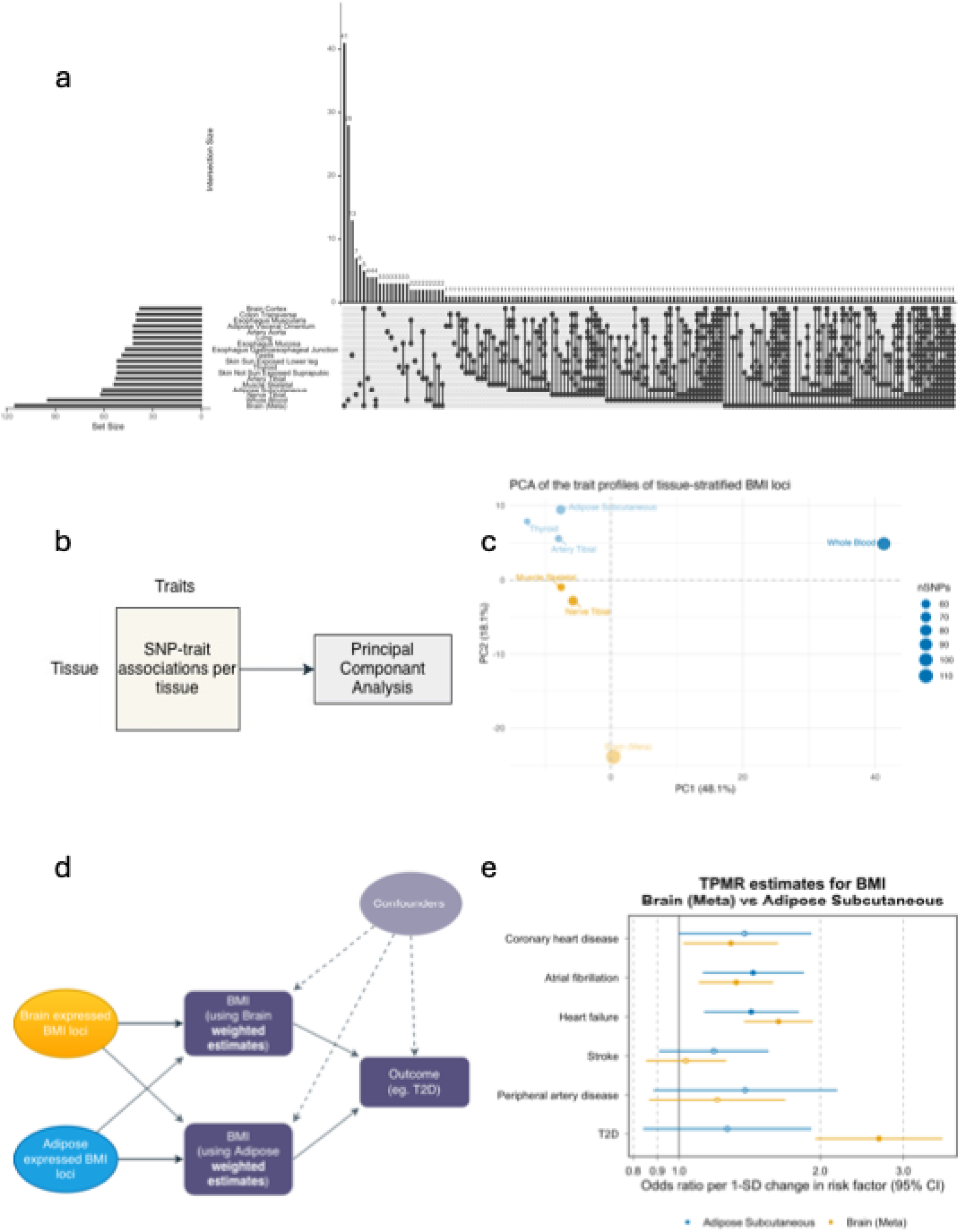
Tissue-informed phenotypic refinement of body-mass index a. Upset plot illustrating sets of BMI colocalization group variants shared across 18 tissues. b. Outline of principal component analysis (PCA) applied to evaluate the composition of trait associations per tissue. c. Results of PCA analysis, with PC1 and PC2 plotted along the x and y axes respectively. d. Directed acyclic graph (DAG) outlining tissue-stratified multivariable Mendelian randomization (MR) analysis. e. The results of tissue-stratified multivariable MR analysis evaluating the direct effects of genetically predicted BMI variants sharing colocalization groups with subcutaneous adipose and brain tissue on cardiovascular and cardiometabolic outcomes.

**Figure 6:**
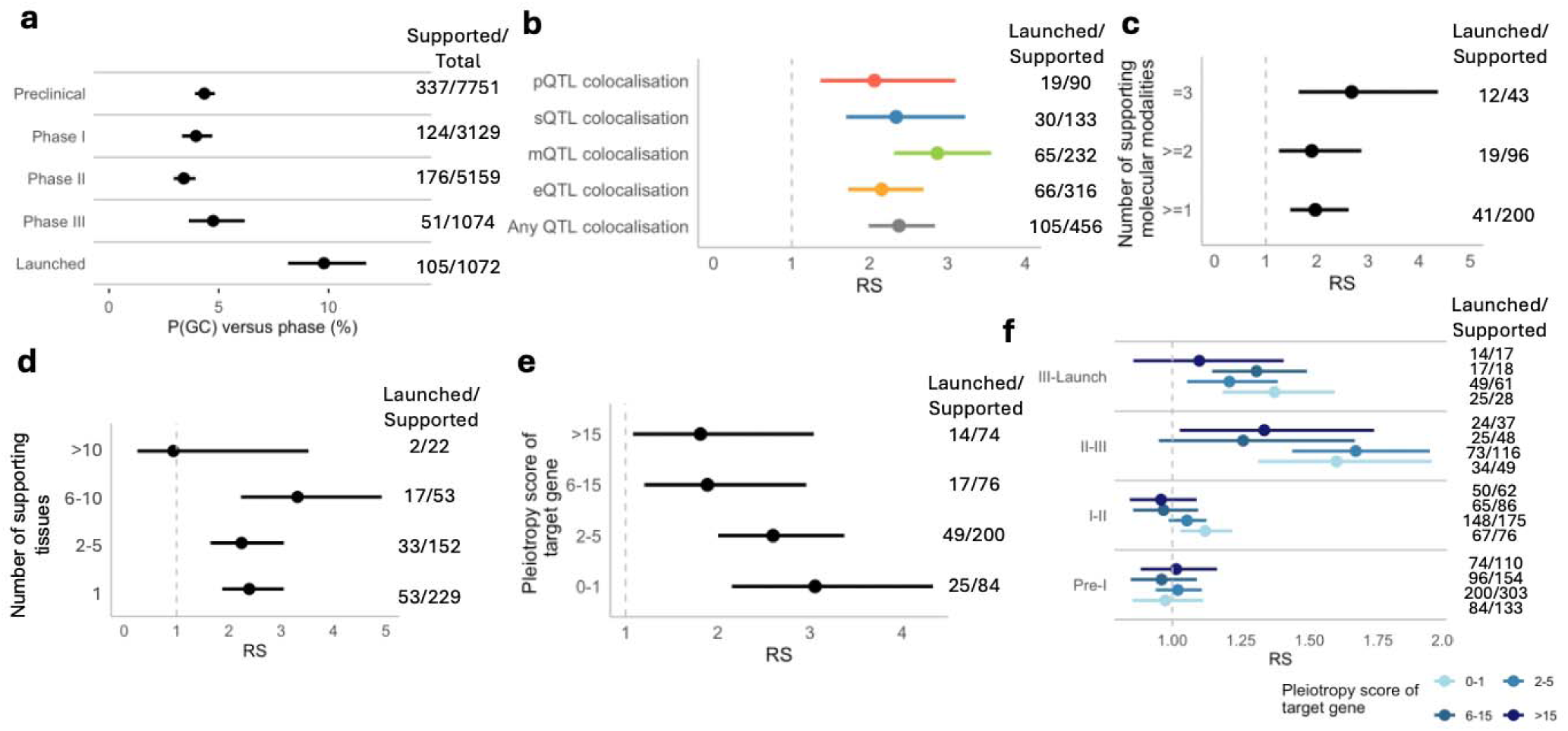
Genetic colocalization evidence improves drug target success. **a**, Proportion of T-I pairs (active and historical combined) with genetic colocalization support P(GC), split by maximum clinical development phase reached. Values to the right indicate the number of GC supported/total T-I pairs per phase. **b**, Overall relative success (RS; phase I–launched) of T-I pairs with GC support, split by molecular mode of support. Value right indicate the number of T-I pairs with the indicated mode of support (denominator) which were launched (numerator). **c**, Overall RS of T-I pairs split by the number of modes of supporting molecular evidence (eQTL, mQTL or sQTL), restric to drug target genes with three modes of genetic insight available. **d**, Overall RS of T-I pairs split by the number of tissue types showing genetic evidence. **e**, Overall RS of T-I pairs split by the pleiotropy score of the target gene and **f**, split by the phase of drug development. For c-f right hand side values are as in b. Error bars represent 95% confide intervals (Wilson’s for P(GC) and Wald for RS).

We examined the complex trait profile for BMI colocalization groups, stratified by tissue annotation. Loci with colocalization between whole blood QTLs and BMI showed the largest number of colocalizations with other complex traits (407 unique complex trait signals across 95 loci), with brain-annotated loci comprising the next largest (246 complex traits across 115 loci; **Supplementary Table 8**). We built a tissue-by-trait matrix of the number of BMI loci shared, and principal component analysis (PCA) was applied to determine how distinct the pleiotropy patterns were by tissue (see **Methods** for detail) (**Figure 6b & 6c**). The results of PCA analysis indicated strong separation of whole blood from other tissues along PC1 (48.1% of variance).

Brain tissue and subcutaneous adipose tissue were separated along PC2. Further characterization of trait enrichment by tissue indicated that subcutaneous adipose tissue loci were enriched with lipid-ratio measures, brain tissue loci were enriched for cardiovascular and lifestyle traits, whereas whole blood tissue loci exhibited pleiotropy with a broad set of systemic measures and was generally more heterogeneous than other tissues (**Supplementary Note 5**).

We aimed to demonstrate whether BMI loci stratified by tissue annotations contributed differentially to the effect of BMI on secondary cardiovascular and cardiometabolic outcomes as demonstrated previously in the literature using a tissue-partitioned multivariable MR approach(30). In these studies, differential effects were observed for subsets of BMI associated SNPs identified by evidence for colocalization in subcutaneous adipose and brain tissue when incorporated as separate exposures in a multivariable MR framework (**Figure 6e**). In the present study, ‘brain-BMI’ and ‘adipose-BMI’ exposures were derived by weighting the SNPs with evidence for colocalization (H_4_≥0.8) in either tissue by their tissue specific H_4_ value, or down-weighting estimates towards the null if they did not reach the threshold for colocalization. The results of multivariable MR analyses estimating the direct effects of brain-and adipose-BMI in the present analysis provided strong evidence of a differential effect of brain-BMI on type-2 diabetes when accounting for the effect of adipose in the model (brain-BMI: Odds ratio (OR)= 2.66; 95% confidence-interval (CI)= 1.95-3.63, P=5.79e-10; adipose-BMI: OR=1.23, 95%-CI=0.84-1.91, P=0.23), replicating the finding described previously (31). However, similar differential effects on cardiovascular outcomes such as coronary heart disease and atrial fibrillation did not replicate in the present study likely due to differential power in the input GWAS for BMI to yield shared tissue signals compared to the original analysis (**Figure 6f**, **Supplementary Table 9**).

### Colocalization, tissue specificity and pleiotropic effects contribute to clinical trial success

To test the utility of the GPMap in identifying actionable biological links between molecular perturbations and disease states, we assessed whether drug target genetic colocalization (GC) support improved the likelihood of drug success. We followed the approach presented by Minikel et al.(32) utilising 18,185 historical and active drug target-disease indication (T-I) pairs for which both the molecular target and the disease indication matched at least one GPMap trait (**see Methods** for trait mapping; totalling 453 unique disease indications and 2221 unique drug target genes; **Supplementary Table 10**). Of the drug targets with available molecular data in the GPMap, 99.3% had eQTL, 92.6% sQTL, 88.9% methQTL and 38.0% pQTL associations. GC support was given if both a target QTL and disease indication mapped trait appeared in at least one colocalization group.

The proportion of T-I pairs with colocalization support, P(GC), was 2.4 times higher for launched drugs than for those that failed at any phase during clinical development (**Figure 5a**). Notably, this was substantially higher than if only pairwise colocalization evidence was considered (P(GC) = 1.7), indicating the strength of trait clustering to rescue false negative colocalization results. We defined overall success P(S) as the proportion of drugs entering Phase I that were subsequently launched. Relative success (RS) was taken as the ratio of the probability of success with:without GC support. P(S) for a given T-I pair was 2.4 [95% CI 2.0-2.8] times higher if supported by colocalization of at least one variant from the disease indication GWAS with any target QTL, and highest if it possessed methQTL support (RS=2.9 [2.3-3.6]; **Figure 5b**). Of the 744 targets that had gene expression, gene splicing and methylation level insight, RS was 35% higher if the target exhibited GC across all 3 (RS=2.7 [1.6-4.4]) relative to any combination of one or more (RS=2.0 [1.5-2.6]; **Figure 5c**).

We next assessed whether the tissue specificity of a drug target gene associated with the likelihood of a drug candidate progressing through clinical pipelines. RS was greatest (>2) for those targets demonstrating GC support in a restricted set of tissues, with those showing low tissue specificity (colocalizing QTLs observed across >10 tissues) being no more successful than targets lacking GC support (RS=0.93 [0.25-3.5], **Figure 5d**).

We hypothesised that targeting highly pleiotropic genes that are involved in diverse molecular pathways may increase risk of off target effects that halt drug development. Drugs targeting genes that exhibit low pleiotropy (score of 0-1; i.e. QTLs for these genes exhibit, at most, QTL associations with one other gene, see **Methods**) demonstrated an overall RS 1.7 times higher than highly pleiotropic genes (score of >15; RS=3.1 [2.2-4.3] compared to RS=1.8 [1.1-3.0] **Figure 5e**). When split across clinical development phases, pleiotropy profile most strongly differentiated the RS of drug candidates transitioning from Phases II-III (**Figure 5f**). Overall, these analyses validate GPMap reported genetic colocalizations, which capture relevant genetic links between molecular perturbations and disease states that underly successful drug development programmes. Characterisation of the tissue and pleiotropy profiles of drug target genes may aid future target prioritisation and prediction of success across developmental phases.

## Discussion

GWAS have now been conducted for an expansive set of human phenotypes, from molecular measures to complex traits and diseases. Our study aims to build on this wealth of information by organizing shared genetic factors across the phenome into a putative GPMap. It pursues the hypothesis that leveraging comprehensive maps of pleiotropy will inform mechanistic insight into disease aetiology. The resource has been made available through an accessible analytical platform, and we have demonstrated its utility here for interrogations into the fundamental nature of genetic pleiotropy, deconstructing trait-level mechanisms, and informing drug target prioritization. This resource currently uses >2.5 million complex and molecular traits, but this will expand as more data becomes available. Our platform also provides a tool for GWAS to be uploaded to generate a bespoke GPMap colocalizing that trait against all others.

Existing strategies to functionally annotate GWAS loci, including deriving variant deleteriousness scores(33), integrating chromatin state maps, and overlaying gene QTL associations(34), can generate extensive candidate gene lists. However, these approaches are unable to resolve the pathways through which a genetic effect is most likely propagated from the coincidental proximity of genomic features. The GPMap overcomes these limitations by integrating millions of fine-mapped and colocalized associations, enabling the empirical mapping of gene-trait relationships while explicitly accounting for LD-induced confounding.

We demonstrate the strengths of this resource for both locus and trait-level deconvolution. The colocalization of several tiers of molecular QTLs, physiological measures and health outcomes at loci associated with haemoglobin levels enables conclusive gene tagging, and highlights the pleiotropic effects yielded by single variants across distinct biological pathways. As well as improving understanding of gene function, knowledge of pleiotropy at the gene level allows the potential consequences of directly interfering with a gene product to be assessed. Evidencing the utility of this, we demonstrate that quantified genetic pleiotropy, as well as the tissue specificity of drug target genes, associates with drug success. This knowledge can help inform drug target prioritization by exposing potential off-target effects, as well as exposing drug repurposing opportunities for diseases with shared genetic aetiology. Furthermore, the GPMap enables the systematic identification of’trait-exclusive’ loci, genes or variants exhibiting minimal pleiotropy. These high-specificity signals represent promising therapeutic targets. Such variants serve as ideal instrumental variables for Mendelian Randomization (MR) studies, providing the biological clarity required to streamline drug development pipelines and prioritize targets with the greatest translational potential.

The genetic variants associated with complex traits are likely to exert their effects on that trait via distinct biological processes. In the context of MR, partitioning IVs by evidence for genetic colocalization with relevant tissues has proven informative in identifying examples where the overall exposure effect on an outcome is differentially influenced via a particular pathway or tissue(27–29). Key to the derivation of partitioned exposures is that sets of IVs reliably capture differential biological effects. We demonstrate here how the GPMap can facilitate the identification of IVs (e.g. for BMI) which are functionally linked to a tissue (brain-BMI, adipose-BMI), and which can be further characterized based on the pleiotropy profile of their respective CGs. The identification of refined phenotypic effects can help determine the disease context of a particular mechanism, offering unique insight for novel targeted interventions.

While the GPMap provides a systematic high-resolution atlas of genetic pleiotropy, its interpretation is bounded by two primary limitation categories. The first encompasses constraints inherent to the input GWAS summary statistics and the sensitivity of the underlying colocalization and fine-mapping algorithms, which are detailed in **Supplementary Note 6**. The second category relates to the structural and methodological parameters established during the construction of the GPMap itself.

Chief among these is that our current map represents a lower bound of global pleiotropic architecture. Both statistical power and phenotypic coverage act as primary bottlenecks; variants with modest effect sizes or those influencing traits not captured in current biobanks will inevitably result in an underestimation of shared genetic signals. Consistent with previous findings, we observed a correlation between the power of the underlying summary statistics and the consistency of colocalization results(35), which provided inconsistencies when clustering.

The trait categorization schema used to inform trait level pleiotropy scores, while designed to provide biologically interpretable annotations, introduces a degree of heuristic classification. Our pleiotropy scores are necessarily sensitive to these categorical boundaries, and alternative taxonomies of human disease could shift the perceived breadth of a gene’s influence.

Finally, a significant limitation of the current GPMap is its reliance on data derived from individuals of European ancestry, borne from the relative scarcity of large-scale GWAS and QTL datasets for non-European ancestries. Because LD structures and allele frequencies vary across populations, the pleiotropy identified may not perfectly translate across diverse ancestral backgrounds. Expanding the GPMap to include multi-ancestry proteomic and transcriptomic data remains a critical frontier for improving fine-mapping accuracy and ensuring the global portability of this resource.

While the observed colocalizations provide strong statistical support for shared genetic mechanisms, functional characterization remains essential to definitively establish the causal links between the identified genes and the complex trait. It is important to note that the identified candidate variants represent the statistically most probable causal candidates across the integrated traits rather than confirmed functional drivers. Nonetheless, the systematic tagging of these variants to specific genes through shared QTL signals, rather than mere genomic proximity, significantly enhances their utility.

The GPMap will be continuously updated with available and emerging complex and molecular traits to provide a deeper understanding of genotype to phenotype relationships. This will including complex traits with larger sample sizes from population cohorts and disease-focused consortia, chromatin accessibility studies and trans-QTL markers. Trans markers, which show associations with distal genes, enable a deeper understanding of genome regulation, including the relevance of non-coding regions, as well as assessment of horizontal pleiotropy and reverse causal effects. The map will also benefit from including additional data from alternative sources, namely experimental gene perturbation data and combining it with GWAS summary statistics(36), which will allow for a deeper ability to understand the biological mechanisms of pleiotropy.

Overall, we provide an open-source resource that integrates GWAS insight from across the phenome to empower gene and trait-centric interrogations into the complexity of human biology. We demonstrate that informing downstream analysis with comprehensive empirical measures of pleiotropy can substantially enrich mechanistic understanding, and has powerful, far-reaching applications.

## Methods

Figure 1 represents an overview of the pipeline steps in building the GPMap, and an overview of the software architecture is available in **Supplementary Note 7**. The data processing pipeline was orchestrated by Snakemake(37), the code is open source and can be viewed in the GitHub repository https://github.com/MRCIEU/genotype-phenotype-map. Further elaboration of definitions of the subsequent terms is available in **Supplementary Note 1**. A set of definitions for data extraction is available in **Supplementary Table 11**.

### Data Collection, Extraction, and Curation

We aggregated summary statistics from multiple resources to capture diverse molecular and phenotypic associations. Complex trait associations were sourced from the EBI GWAS Catalog(38) and OpenGWAS(39) UK Biobank studies, supplemented with rare variant findings from GeneBass(40), Backman et al.(41) and AstraZeneca Genomics Initiative(42), which also includes rare variant plasma protein level associations from UK Biobank. Molecular QTL data includes all *cis* Gene Expression Quantitative Trait Loci (eQTL) and Splice Variant Quantitative Trait Loci (sQTL) from GTEx v10(43) across 49 tissues, eQTLGen(44) *cis* and *trans* eQTLs, *cis* Protein Quantitative Trait Loci (pQTL) from the UK Biobank Pharma Proteomics Project(45), DNA Methylation Quantative Trait Loci (methQTL) from GoDMC(46), and single-cell eQTL data from OneK1K(47). Brain-specific eQTL associations were derived from BrainMeta(48) and Brain-eMeta(49) resources.

### LD Partitioning

Analysis was restricted to autosomes (chromosomes 1-22), partitioned into 1,357 linkage disequilibrium (LD) independent regions using LDetect boundaries based on the European labelled sample subset in 1000 genomes (50) for GRCh38 reference build. An ancestry-matched LD reference panel was constructed from 50,000 randomly selected UK Biobank individuals(51) with European ancestry using variants imputed against the TOPMED reference panel. Variants with imputation quality scores (r²) < 0.8 or minor allele frequency (MAF) < 0.5% were excluded.

### Data Standardization and Imputation

All QTL datasets were converted to the Binary Effect Size Distribution (BESD) format using the SMR toolkit (52) to enable standardized processing. For all traits (including molecular traits in BESD format), for each LD partition we extracted the summary statistics regions that exhibited at least one association at p<1.5e-4, which were then converted into a standard format and harmonized(53). Harmonization involved creating a unique SNP id that would be matched across all studies, in the format of “CHR:BP_A1_A2”, where A1 is chosen in alphabetical order, and other fields (BETA, EAF) were altered accordingly. Before reporting, all SNP IDs and effect estimates were standardised to the REF/ALT allele calling convention used by NCBI dbSNP (effect estimates corresponding to each gain of an ALT allele). If allele frequency data were missing, we used allele frequencies from the reference panel. Only SNPs that were in the reference panel were retained. If there were fewer than 150 SNPs in the LD region and the input data was not sparse or a rare variant trait (**Supplementary Note 1**), that trait region was discarded.

We then performed imputation on the summary statistics to fill the missing data which arose due differing genotyping chips across studies. Existing summary statistic imputation methods (ssimp(54), RAISS(55)) provided significant computational constraints, therefore we developed an approximation that maintains accuracy while increasing computational performance by over 900% (**Supplementary Note 8**).

### Statistical Fine-mapping and Quality Control

Recent studies have demonstrated that colocalization combined with fine-mapping improve on other approaches to identify shared causal variants(56,57). We performed probabilistic fine-mapping of each LD block independently using SuSiE(12) with default parameters (prior variance = 50, maximum 10 credible sets per region). Regions where SuSiE failed to converge or identified a single credible set retained their original summary statistics. For regions with >3 credible sets, we applied DENTIST(58) to identify and remove variants showing discordance between observed and LD-imputed association statistics. If DENTIST identified and removed SNPs that did not pass quality control steps, those SNPs were removed, and the fine-mapping step was rerun.

We observed instances where the SuSiE model yielded inflated LBF scores. Specifically, these scores achieved statistical significance despite the corresponding marginal association data showing no such evidence. To address this discrepancy, an additional post-hoc quality control (QC) filter was implemented. This filter involved calculating an approximate p-value from the SuSiE-derived LBF score. We then compared this derived p-value against the original marginal p-value. A SNP was designated as problematic and subsequently excluded from the LBF results if the original marginal p-value exceeded a threshold of 1e-3, while the LBF-derived p-value was simultaneously below 1.5e-4.

Although it is possible for variants to gain significance after accounting for independent signals in LD with opposing directions of effect, we found in the majority of these cases the genotyped signal was unsupported by proximal variants in the region. These were likely artifactual signals arising due to a mismatch between the study population and the LD reference panel used for fine-mapping.

### Pairwise Colocalization

Harmonized summary statistics were grouped by genomic position (within 50Kb) and each pair was analysed using coloc(59) to assess evidence for shared causal variants across traits. Only loci with ≥50 overlapping SNPs were considered. Instead of comparing every extracted and fine-mapped result in an LD block with to every other, only extractions where fine-mapping had selected a SNP in the credible set less than 50 kilobases from other fine-mapped results were included for coloc analysis.

### Graph-based clustering of colocalization results into Colocalization Groups

After pairwise colocalization calculations were run, the results were clustered into colocalization groups using a two-step process, where a vertex is a trait which has a minimum p-value below p<1.5e-4, and an edge is a colocalization result of H_4_≥0.8 between two traits. First, we used the infomap algorithm(60) available from the R package igraph(61) to cluster the results. Second, we removed all nodes from the cluster that had a <5% amount of connectivity compared to the most connected vertex in the cluster. Finally, we removed both the edges that were found to be between clusters and orphaned vertices after the edge pruning. Further simulations, and explanations on the justification for these thresholds is available in **Supplementary Notes 2-3**.

Each remaining colocalization group is then assigned a ‘candidate variant’. The variant is chosen by using the log-bayes factor score (LBF) calculated by SuSiE(12). The LBF is a representation of the likelihood that the variant in question is the causal variant. All LBF scores for each SNP in colocalization group are summed, and the SNP with the maximum cumulative LBF score is chosen as the candidate variant.

### Rare Variant Grouping

Analysis for rare variants necessitated a modified pipeline due to practical constraints in data preparation. Specifically, the absence of comprehensive genomic regions surrounding the significant SNPs, combined with the lack of rare variants within our LD reference panel, precluded the use of imputation, fine-mapping, and colocalization methodologies. Instead, we amalgamated all rare variant traits that shared a significant SNP and grouped them based on the shared association signal. Consequently, the resulting rare variant groups are defined exclusively by their shared statistical association. Most rare variant datasets already assigned genes to each SNP via functional annotation. This was kept and reflected in GPMap and exposed as a ‘situated gene’.

### Functional Annotation and Classification

Variants were annotated using Ensembl Variant Effect Predictor (VEP)(62). All molecular QTLs that provided gene and tissue annotation were used, and no other steps to associate a QTL with a gene were taken. Most data supplied from rare variant analyses had pre-defined functional annotation for each variant. Those annotations were preserved and stored under the name ‘situated gene’ in the rare results.

Gene annotation was provided from Ensembl BioMarts(63). Each QTL resource has annotated each measurement with either a gene name or ENSG id. We queried bioMart hsapiens_gene_ensembl database to annotate each gene using the gene symbol where possible.

### Trait categorization

Traits were categorized into 23 categories: infectious disease, neoplasm, disease of blood and blood-forming organs, metabolic disease, neurological disease, mental disorder, nervous system disorders, disease of eye and adnexa, disease of ear and mastoid process, disease of circulatory system, disease of digestive system, disease of skin and subcutaneous tissue, disease of musculoskeletal system and connective tissue, disease of genitourinary system, physiological measures, reproductive measures, psychiatric measures, anthropometric measures, environmental measures, socioeconomic measures, behavioral measures, cell traits, targeted protein measure.

This was achieved by generating a prompt for OpenAI model gpt-5-nano for each complex trait analyzed (excluding molecular phenotypes) requesting the best matching category for each trait name along with a confidence score for the match. Trait to category mapping was manually inspected. A number of traits had a low category match confidence score but were retained for completeness. Some traits were too broad and therefore were manually set to undefined (See **Supplementary Note 1**).

The differentiation between a highly specific measurement and a QTL pose problems, hence 2 of the 23 trait categories have a unique standing, ‘Cell Traits’ (eg. ‘IgD-CD27-B cell %B cell’) and ‘Targeted Protein Measure’ (eg. ‘VDBP plasma levels’) are neither considered a complex trait nor a molecular trait.

### Pleiotropy Score Calculation

We used two distinct metrics to quantify pleiotropy: trait category pleiotropy and gene pleiotropy, and these were each calculated at both the gene level and the variant level. For variant level gene pleiotropy, we counted how many distinct gene-based molecular traits colocalised with that variant. For variant-level trait category pleiotropy we counted how many trait categories colocalised with the variant. Both were based on colocalization at the CG level.

A similar process was followed to obtain a gene pleiotropy score, counting how many distinct gene-based molecular traits or trait categories colocalised with a molecular trait for the target gene. Rare variant results were excluded from these scores. For all results related to pleiotropy, only genes that were explicitly measured by the QTL resource were used, as such the methQTL results were excluded due to the genes being defined via functional annotation.

### Amalgamating and Publishing Results

Once all data were processed, the data were turned into a series of normalized DuckDB databases to ensure consistency and allow for easy accessibility. These database files are then queried and exposed via an API and an R package https://github.com/MRCIEU/gpmapr. Further data, including all summary statistics are available in an open-access requester pays cloud storage bucket https://console.cloud.google.com/storage/browser/genotype-phenotype-map.

Further, a series of images in SVG format were created to aid with visualization of the results, which are available when accessing the data from https://gpmap.opengwas.io.

The GWAS upload process uses the same code pipeline, packaged into a Docker container and run on a server.

All subsequent analyses of the GPMap were performed using custom R scripts, which have been made publicly available to ensure reproducibility and are hosted in a GitHub repository (https://github.com/MRCIEU/genotype-phenotype-map-analysis/tree/main/scripts/analysis/).

### Haemoglobin case study

Summary statistics for the UK Biobank (UKB) trait, haemoglobin concentration (referred to as the “anchor trait” in this analysis, field ID 30020), were taken from both common variant GWAS(OpenGWAS: ukb-d-30020-irnt)(39) and WES based analyses(41) and extracted into the GPMap under trait ID 4759. Common variant colocalization results (groups of traits sharing a colocalizing signal at anchor trait associated variants and linked through trait clustering) were derived from running the fine-mapping and colocalization pipeline using default parameters as described above, filtering to those regions containing an anchor trait signal at p<5e-8. Regions from colocalizing traits were excluded if the minimum association p-value in that region was >5e-8. Shared rare variant results were taken as WES variants at <1% minor allele frequency that showed an association with the anchor trait (at GWS) and the trait. Colocalization/rare variant sharing results were accessed from the GPMap API using the gpmapr package trait()function.

Traits deemed to be assessing highly correlated or duplicate UKB traits (mean corpuscular haemoglobin, mean corpuscular haemoglobin concentration, glycated haemoglobin HbA1c) were excluded from downstream analyses. Trait clustering was performed on all phenotypic traits showing >1 colocalizing genetic signal with the anchor trait. Duplicate traits were crudely removed if the trait name was identical, retaining the study with the largest sample size. A trait-by-trait matrix was generated, enumerating the number of independent genetic variants genome wide that colocalized between the two traits and the anchor trait (i.e. a value of 2 would indicate two variants genome wide were determined to be shared by trait 1, trait 2 and haemoglobin concentration following fine-mapping, colocalization and trait clustering). Hierarchical clustering of traits was performed by using hclust() on the distance matrix derived using the dist() function from the stats base R package. For two example loci containing colocalizing signals across both complex traits and molecular traits, *TMPRSS6* and *HP/HPR*, betas were taken from the original reported summary statistics for each trait, or imputed values if necessary, pre fine-mapping.

### Tissue-informed phenotypic refinement analysis

BMI (https://gpmap.opengwas.io/trait.html?id=1992) was queried using the *gpmapr* R package for which 992 independent loci have been assigned to colocalization groups. Approximately a third of BMI colocalization groups (320/992) were shared with any tissue-derived eQTL. A summary of the number of loci attributed to each tissue-type is provided (**Supplementary Table 9)**.

To assess whether tissue eQTL annotations elucidate distinct biological effects underlying BMI, we evaluated the complex trait profile of sets of loci annotated by tissues with the largest tissue-specific contribution and measured in both sexes (subcutaneous adipose, tibial artery, brain, skeletal muscle, tibial nerve, thyroid and whole blood) (Figure 6a). First, tissue annotated BMI colocalization groups were evaluated by principal component analysis (PCA) on the relative representation of traits per tissue. Briefly, we identified the set of all complex traits present within BMI colocalization groups. PCA was applied to a trait-standardized tissue-by-trait variant count matrix to identify between tissue variation in trait representation. Secondly, we applied hierarchical clustering using the *pheatmap* R package to a tissue-by-trait matrix of z-scores derived from the within-tissue proportions of variant–trait associations. Z-score transformation standardized each trait across tissues, enabling comparison of relative enrichment patterns rather than absolute counts.

We demonstrate an applied example of how the GPMap resource may be used to facilitate tissue-partitioned MR analyses using BMI as examplar. This was achieved by incorporating the subsets of genetic variants annotated by tissue in the GPMap as instrumental variables. Specifically, we incorporated the variants annotated by brain and subcutaneous adipose tissue based on the following: functional relevance to BMI, evidence for colocalization group membership specific to each tissue, evidence for differential trait composition in PCA and clustering analyses and to replicate findings attributed to adipose-and brain-BMI by tissue-partitioned MR described in the literature previously. A total of 153 variants were incorporated into a multivariable MR framework (115 annotated by brain (meta), 61 by subcutaneous adipose, 23 shared). ‘Brain-BMI’ and ‘Adipose-BMI’ exposures were derived by weighting the SNPs with evidence for colocalization (H_4_ ≥ 0.8) in either tissue by their tissue specific H_4_ value, or down-weighting estimates towards the null if they did not reach the threshold for colocalization as described. Conditional F-statistics remained >10 for each tissue-annotated exposure variable, indicating instrument strength in the multivariable framework.

### Drug target – disease indication pairs analysis

We utilised 25,713 active and historical drug target-disease indication (T-I) pairs with a reported maximum clinical development phase reached as published in Supplementary Table 1 by Minikel et al.(32), derived from the Citeline Pharmaprojects database of drug development programmes prior to December 2022(64). The table of available trait genetic associations that could be mapped to a corresponding target gene (sourced from OMIM, OTG, PICCOLO, Genebass and IntOGen) was taken from the public data repository(64) and used to derive a reference list of association study trait to MeSH ID and MeSH term mappings. Extracted GPMap complex traits were matched to this reference list of trait names using ChatGPTv5, prompted to provide the best matching or most closely related reference trait for each GPMap trait, and provide a confidence score between 0 and 1. Pairings were manually checked and any GPMAP trait that was not deemed a sufficient match to a reference trait or the MeSH term assigned to it was excluded. The MeSH ID similarity matrix generated by Minikel et al. was used to retain those GPMap traits for which the assigned MeSH term matched a disease indication MeSH term with >=0.8 similarity. If a GPMap trait provided a high similarity match to multiple disease indication MeSH terms all pairings were retained. GPMap molecular traits were filtered to those assaying the transcript or protein expression or alternate splicing of a drug target gene, or methylation of a CpG site mapped to a drug target gene, resulting in a total of 18,932 of the original 25,713 T-I pairs available (74%) with GPMap insight. Disease indication traits that did not appear in any colocalization clusters due to weak regional signals were excluded, resulting in a final set of 18,185 T-I pairs for analysis (including 1,211 GPMap complex traits falling under 305 unique MeSH IDs and mapped to 453 unique disease indication MeSH IDs with similarity of >=0.8, and paired with 2221 drug targets assayed in molecular studies (S**upplementary Table 10**).

The colocalization results for all disease indication traits were extracted from the GPMap using the gpmapr package. A T-I pair was taken to have genetic colocalization (GC) support if at least one indication associated variant and a drug target QTL (at p<5e-8) fell within the same colocalization cluster. If the indication MeSH term mapped to multiple studied traits, GC support could be contributed by any of these. The proportion of T-I pairs with genetic colocalization support P(GC) was taken as the number of pairs with GC support over the total number of pairs with GPMap insight, split by maximum phase of development (preclinical, phase I, phase II, phase III or launched). Success at a given phase was taken as a drug proceeding to the next stage in the development pipeline, and relative success (RS) was taken as the proportion of successful drugs (T-I pairs) with GC support divided by the proportion successful without GC support.

Overall RS was calculated for those drugs which entered phase I trials and were subsequently launched (e.g. excluding programmes that failed at the preclinical phase). Overall RS was also calculated across subgroups of drug targets, split according to the type and number of supporting molecular modalities (pQTL, sQTL, methQTL and eQTL evidence), the number of tissues contributing genetic support, and the pleiotropy score of the target gene (calculated as described above). Whole blood and tissue specific eQTLs and sQTLs were taken from GTEx, eQTLs from Brain eMeta/Meta, pQTLs from the UK Biobank (plasma only) and methQTLs from GoDMC (whole blood only). Where possible, tissues were grouped into categories: brain/nervous system (brain and nerve), blood cell (peripheral blood mononuclear cells, cells and blood), kidney (adrenal and kidney), heart (heart and artery) and digestive (saliva, esophagus, colon, intestine and stomach).

### Open-Source Code for Methods

The code associated with each step described above can be found in the GitHub repository https://github.com/MRCIEU/genotype-phenotype-map, under the pipeline-steps directory. Each step is invoked as a script from the Snakefile in the same repository.

## Data and Code Availability

We have provided multiple ways for users to use and interact with the data.

For an interactive exploration of the data, please visit https://gpmap.opengwas.io/. To upload a GWAS that will be run against all existing data in the database, either use the website (https://gpmap.opengwas.io?showUpload=true) or use the R package gpmapr (https://github.com/MRCIEU/gpmapr). Vignettes have been created to illustrate how to use the R package (https://mrcieu.r-universe.dev/gpmapr).

We have also published the database of results, along with all fine-mapped summary statistics in a cloud storage bucket. As the cost to hosting this amount of data is high, we have set this to a ‘requester pays’ model, where the user pays for the cost to download. The bucket is available at https://console.cloud.google.com/storage/browser/genotype-phenotype-map.

The code for this project is available over 3 different publicly available GitHub repositories. The data processing pipeline is available at https://github.com/MRCIEU/genotype-phenotype-map, the website and API are available at https://github.com/MRCIEU/genotype-phenotype-api, and the R package is available at https://github.com/MRCIEU/gpmapr. As the data is large and available via both the R package and website, we have not included the results as supplementary material.

The data from the original studies is available via the R package, *source_url* is provided for each colocalization result.

## Acknowledgements

We express our sincere gratitude to the research participants and the investigators of the various studies and consortia whose data contributed to the development of the GPMap. This work would not have been possible without the altruistic contribution of hundreds of thousands of individuals to genetic research.

We specifically acknowledge the following biobanks and consortia for providing the GWAS summary statistics and functional genomic data: UK Biobank, The GTEx Consortium, FinnGen, and the NHGRI-EBI GWAS Catalog. We also thank the IEU OpenGWAS project for providing the computational infrastructure and data standardization that facilitated this large-scale integration.

## Contributions

**ARE**: methodology, software (data processing, web application, R package), validation, formal analysis, investigation, data curation, visualisation, manuscript draft and editing. **ALH**: methodology, software (data processing), validation, formal analysis, investigation, data curation, visualisation, manuscript draft and editing. **GML**: validation, formal analysis, investigation, data curation, visualisation, manuscript draft and editing. **JJ**: software (web application). **GDS**: discussion, interpretation of the results, manuscript editing. **LP**: funding acquisition, discussion, interpretation of the results, manuscript editing. **TRG**: funding acquisition, discussion, interpretation of the results, manuscript editing. **GH**: funding acquisition, conception, methodology, software (data processing), formal analysis, investigation, manuscript draft and editing. All authors provided relevant input at different stages of the project and approved the final manuscript.

## Additional Information

Supplementary tables and supplementary notes and figures are available for this paper.

