## Supplementary Notes and Figures for "Building The Human Genotype-Phenotype Map to Harness Pleiotropy and Refine Disease Mechanisms"

Supplementary Information

### Supplementary Note 1: Glossary and definitions

**Pleiotropy:**

The definition of pleiotropy varies across academic domains and contexts. For the purposes of this study, we define a distinction between two forms of multi-trait association based on the underlying genetic architecture of a locus:

- **LD-induced Pleiotropy**: Defined as a multi-trait association arising from distinct causal variants located within the same linkage disequilibrium (LD) block. In this scenario, the phenotypic associations are physically proximal but genetically independent.
- **Biological Pleiotropy**: Defined as a multi-trait association driven by a shared causal variant. This indicates a singular genetic origin for the divergent phenotypic effects.

Consistent with this framework, the term ‘pleiotropy’ as used throughout this manuscript refers specifically to biological pleiotropy unless otherwise qualified. Furthermore, we define the task of ‘mapping pleiotropy’ as the systematic effort to distinguish true biological pleiotropy from LD-induced pleiotropy using statistical colocalization and clustering methods.

Further, additional contextual mentions of pleiotropy include:

- **Vertical Pleiotropy**: when a genetic variant affects other traits (which directly mediate the influence on the outcome) via its effect on the initial exposure (e.g. expression of a gene).
- **Horizontal Pleiotropy**: when a genetic variant influences a trait (directly or indirectly, through other traits) independently of the hypothesized exposure.

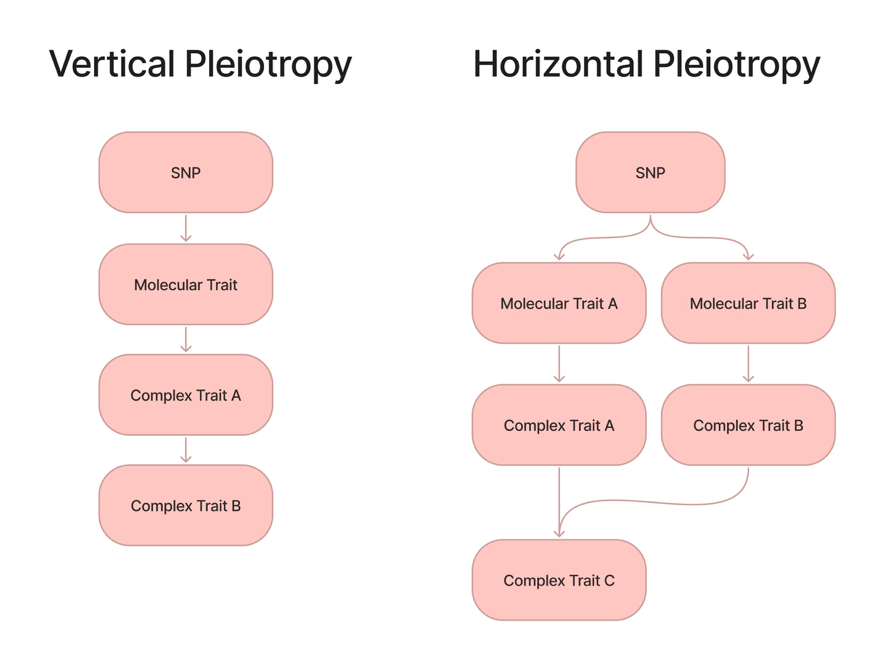

**Figure SN1:** Visual Representation of vertical vs. horizontal pleiotropy

**Pleiotropy score:** There are two pleiotropy scores calculated on both the variant and gene level. The first is the number of distinct trait categories, and the second is the number of distinct protein coding genes, that fall in colocalization groups tagged by the variant (variant level) or containing gene-specific QTLs (gene level). Rare variant results are not included in the calculation.

**Traits:** A trait denotes the outcome variable assessed in any of the GWAS from which we have taken summary statistics for the GPMap. The trait names remain as defined in the original study. Other commonly used names for trait might be ‘phenotype’ or ‘study’.

- Complex Trait: represent polygenic organismal phenotypes and clinical disease states. All complex traits that have genome wide associations with them.
- Molecular Trait: representing discrete cellular processes such as mRNA expression or protein abundance. Molecular traits do not typically have genome wide data associated with them, but rather have significant *cis* (and sometimes *trans*) signals extracted in specific genomic regions, and are associated with a gene and tissue.

A third category of trait, which falls in between these, are ultra-specific measurements which have genome-wide associations, as opposed to *cis-* and *trans-* windows. These have been denoted as ‘Cell Traits’ (eg. ‘IgD- CD27- B cell %B cell’) and ‘Targeted Protein Measure’ (eg. ‘VDBP plasma levels’) are neither considered a complex trait nor a molecular trait.

**Coverage**: dense vs. sparse. Summary statistics that only published results of SNPs that reached a specific p-value threshold are considered sparsely populated, all others are considered densely populated. Sparsely populated summary statistics had their missing values 0-padded and both imputation and fine-mapping steps were skipped.

**Cell Type:** Some QTLs were derived from ‘single cell’ expression assays which derive gene expression measurements from specific cell subsets following single-cell transcriptional profiling. A list of cell types currently in the map are in **Supplementary Table 11**.

**Gene Annotation:** There are two different types of gene annotation that occur for QTLs, ‘gene’ and ‘situated gene’. ‘Gene’ refers to the gene that was assayed and is annotated by the QTL resource itself (eg. GTEx gene expression of TREML2 in blood is annotated as ‘TREML2’). ‘Situated gene’ is currently only applicable for rare variants studies conducted using whole-exome sequencing data and denotes the gene in which the variant is physically situated. Common variant QTL studies have a ‘Gene’, most rare variant phenotypic studies have a ‘Situated Gene’, and rare variant QTL studies have both a ‘Gene’ and a ‘Situated Gene’, which may differ. Genes have been assigned to methQTLs based on the proximity of the assayed CpG site to a gene. These were taken from the Illumina Methylation EPICv2 manifest(1), tagging CpGs by their proximity to gene bodies and promoter regions.

**Colocalization definitions:**

- **Colocalizing Pair**: A colocalization pair is a single colocalization test run between two traits using coloc(2). Two traits are considered to be a pair if H_4_ **≥** 0.8.
- **Colocalization Group**: A set of traits that have been grouped together by a graph-based clustering and pruning methods from the results of pairwise colocalization analysis. A detailed explanation of the clustering and pruning methods can be found in Supplementary Notes 2-3.
- **Group Connectedness Percentage:** The connectedness of the colocalization group is calculated as the total count of the H_4_**≥**0.8 across all colocalization pairs (edges) in the group, divided by the total number of possible edges in the colocalization group with n nodes, *n(n-1)/2*. A higher connectedness means that the colocalization group is more strongly connected.
- **Candidate Variant**: Each colocalization group is assigned a SNP. The SNP is chosen as the variant with the highest cumulative sum of the log bayes factor (LBF), calculated by SuSiE(3), across every trait in the colocalization group In some situations, the candidate variant may not be the variant with the highest LBF for a given study in the colocalization group, or, if the variant was not genotyped or imputed for that study, may be missing. This variant should not be interpreted as the causal variant for traits in the group, it is instead tagging a shared colocalizing signal in the region.

**P-value thresholds:**

- **Genome wide significance (GWS)**: We utilize the standard European genome significant wide p-value threshold of 5e-8 most calculations in the results.
- **Suggestive Significance**: We have extracted and analyzed all loci above a ‘suggestive significance’ of 1.5e-4. As there are approximately 1 million independent loci across the genome (0.05 / 1m = 5e-8), across 3 billion base pairs. Meaning there are approximately 333 independent regions across every 1Mb, 0.05/333 ~= 1.5e-4.

For the resource, both colocalization group and colocalization pair data is available. Users should be aware of their own p-value threshold for multiple testing correction.

**Cis and Trans:** For common variant QTLs, the *cis* window includes any finemapped loci within ±1 Mb of the SNP tagging the QTL, and trans is defined as any region outside of that window. For rare QTLs, any SNP falling directly within the gene that was assayed is considered *cis* (i.e. the ‘gene’ and the ‘situated gene’ match). If this is not the case, the variant is considered to be a *trans* signal for the assayed gene measure.

### Supplementary Note 2: Clustering Method

We aimed to identify groups of traits that all shared the same causal variant based on the results from pairwise colocalization analysis. We assume that sometimes if traits A and B show colocalization at a locus, and traits B and C show colocalization at the same locus, then in general A and C should also colocalize at that locus. However, we expect that this fully transitive relationship would not manifest empirically because of non-zero false positive rates, and incomplete statistical power.

Figure SN2 shows how colocalization networks appear under different scenarios of false positive and false negative rates. In general, clustering appears to be much less affected by lower power than it is by even very modest false positive rates.

We performed simple simulations to examine performance of clustering methods to recapitulate trait groups that share causal variants based on pairwise colocalization results. Simulations varied the number of clusters, the number of traits in each cluster ${m=\sum m}_{l}$, the false negative rate (FNR) of colocalization within a cluster and the false positive rate (FPR) of colocalization between clusters. Cluster sizes were selected based on a cluster imbalance metric, where 0 is equal number of traits per cluster (no imbalance), and 1 is a single trait per cluster except for one cluster which contains all remaining traits (maximally imbalanced).

We trialed several clustering algorithms implemented in the R/igraph R package, including "cluster_louvain", "cluster_infomap", "cluster_walktrap", "cluster_label_prop", "cluster_fast_greedy", "cluster_leiden_modularity". The “cluster_edge_betweenness”, “cluster_spinglass” and “cluster_optimal” methods were also examined but ultimately excluded due to slow run time.

We evaluated clustering performance based on alignment between simulated membership and predicted membership of traits in a cluster based on the true discovery rate and the true positive rate. Figure SN3 illustrates that the infomap algorithm generally performs best across different scenarios in terms of being robust to high FDR and high FNR.

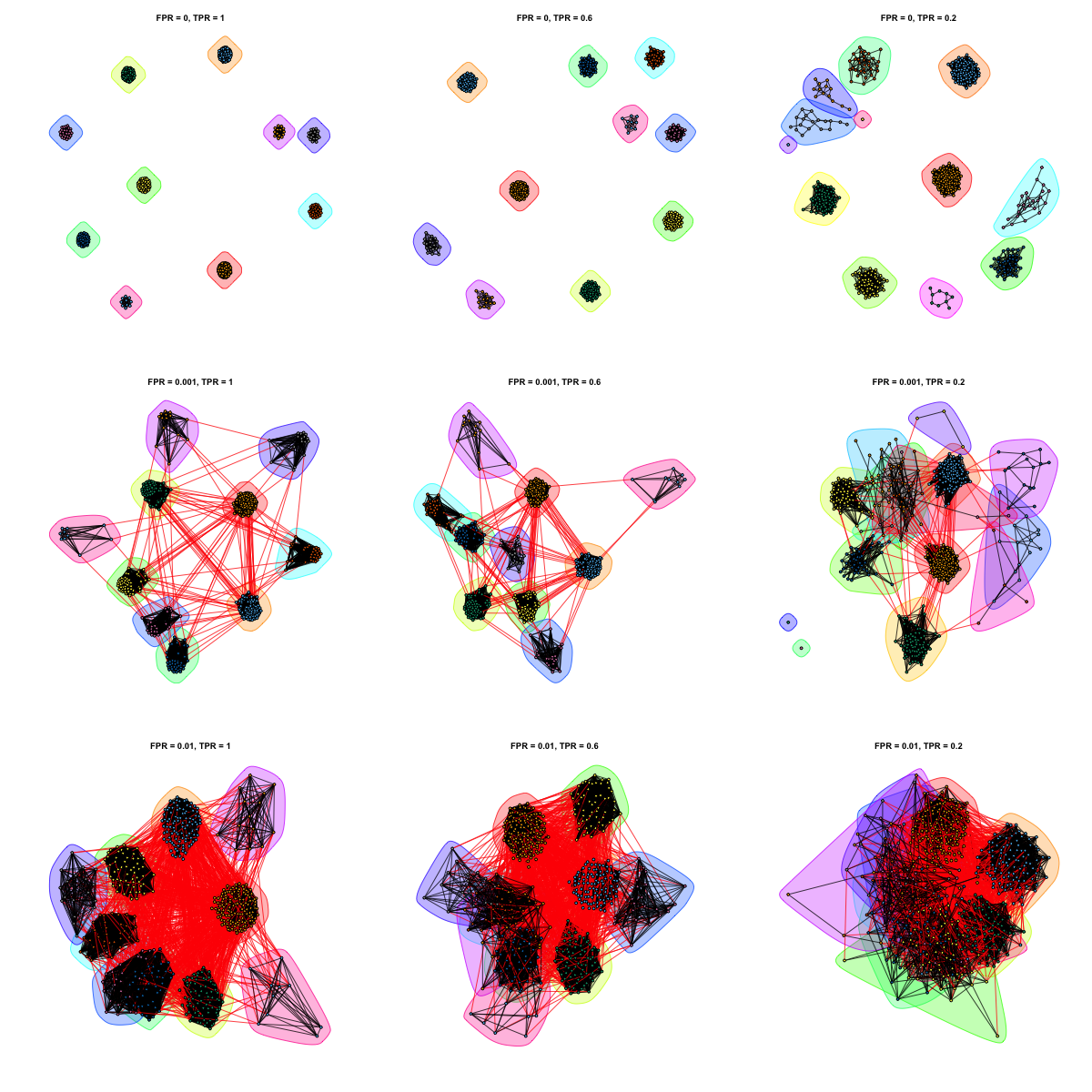

**Figure SN2**: Illustration of clustering method applied to pairwise colocalization

Colocalization networks for 10 causal variants, where the number of traits that share a causal variant will vary across those variants (500 traits in total). Clustering of traits based on their pairwise colocalization is performed using the infomap algorithm in this example. Colors represent inferred colocalization clusters, black lines represent links within inferred clusters and red lines indicate links between inferred clusters. Top left graph is the oracle where the true positive rate = 1 and the false positive rate = 0. Simulated true positive rate (power) decreases across columns, and false positive rate increases across rows.

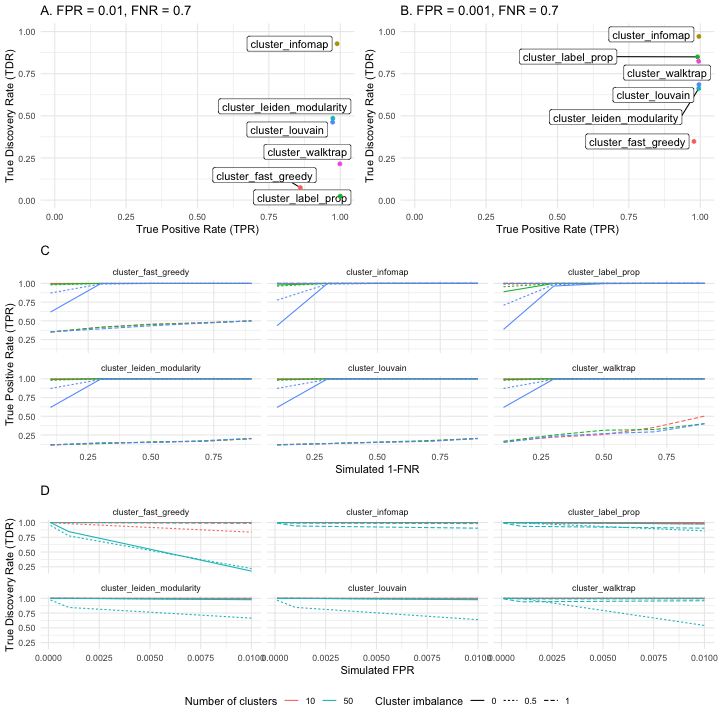

**Figure SN3:** Results of simulations comparing clustering methods

**A**. Example simulation with 2000 nodes, 100 clusters, cluster imbalance = 0.9, FNR = 0.7 and FPR = 0.01. X-axis represents true discovery rates (TDR) of cluster membership, Y-axis represents true positive rates (TPR) of cluster membership for each of the clustering methods tested. **B**. As in (A) but with a lower FPR = 0.001. **C**. Relationship between TPR (y axis) with FNR (x axis) by number of clusters and cluster imbalance for each method. **D.** Relationship between TDR (y axis) and FPR (x axis) by number of clusters and cluster imbalance for each method. Simulation results for (C) and (D) are based on 5 repetitions for each point, and 1000 traits in each simulation.

### Supplementary Note 3: Empirical evaluation of the clustering method

**Colocalization method:**

Prior to performing colocalization of genetic signals across studies (within LD blocks), we performed summary statistic imputation to maximize the overlap of shared variants between studies, followed by fine-mapping using SuSiE(3) to segregate independent signals into fine-mapped credible sets (see **Methods**). We originally trialed identifying groups of studies with a colocalizing fine-mapped genetic signal significant at p<1.5e-4 using HyPrColoc(4), which provides a multi-trait extension to the functionality of coloc(2) to colocalize many hundreds of studies simultaneously. This method reports a single posterior probability (PP) of colocalization and a putative causal variant for groups of studies found to share a distinct colocalizing signal. We found that HyPrColoc was highly sensitive to the number and combination of studies upon which it was run, varying group assignments when new studies were introduced. There was little agreement between the posterior probabilities returned by HyPrColoc for study groupings and the mean pairwise posterior probability of pairwise colocalization (H_4_) between studies in those groups as returned from coloc. Often HyPrColoc assigned high PP (>0.5) to groups of studies with low mean pairwise H_4_, often due to pairs of studies within the group showing greater evidence for H_3_ (independent signals in a region), and vice versa, assigning PP<0.5 to groups of studies showing high pairwise H_4_ (Figure SN4-A). This was also the case for HyPrColoc groups of only two studies, for which we expected higher concordance between methods (Figure S5-B). We did not run a comparison of the two colocalization methods on simulated truth cases to formally assess performance, instead reasoning that reporting coloc pairwise H_4_ calls, alongside the study clusters that were derived based on those calls (as detailed below), would improve the transparency of our results. We anticipated this approach would also reduce volatility in cluster assignments upon integration of new studies into the resource.

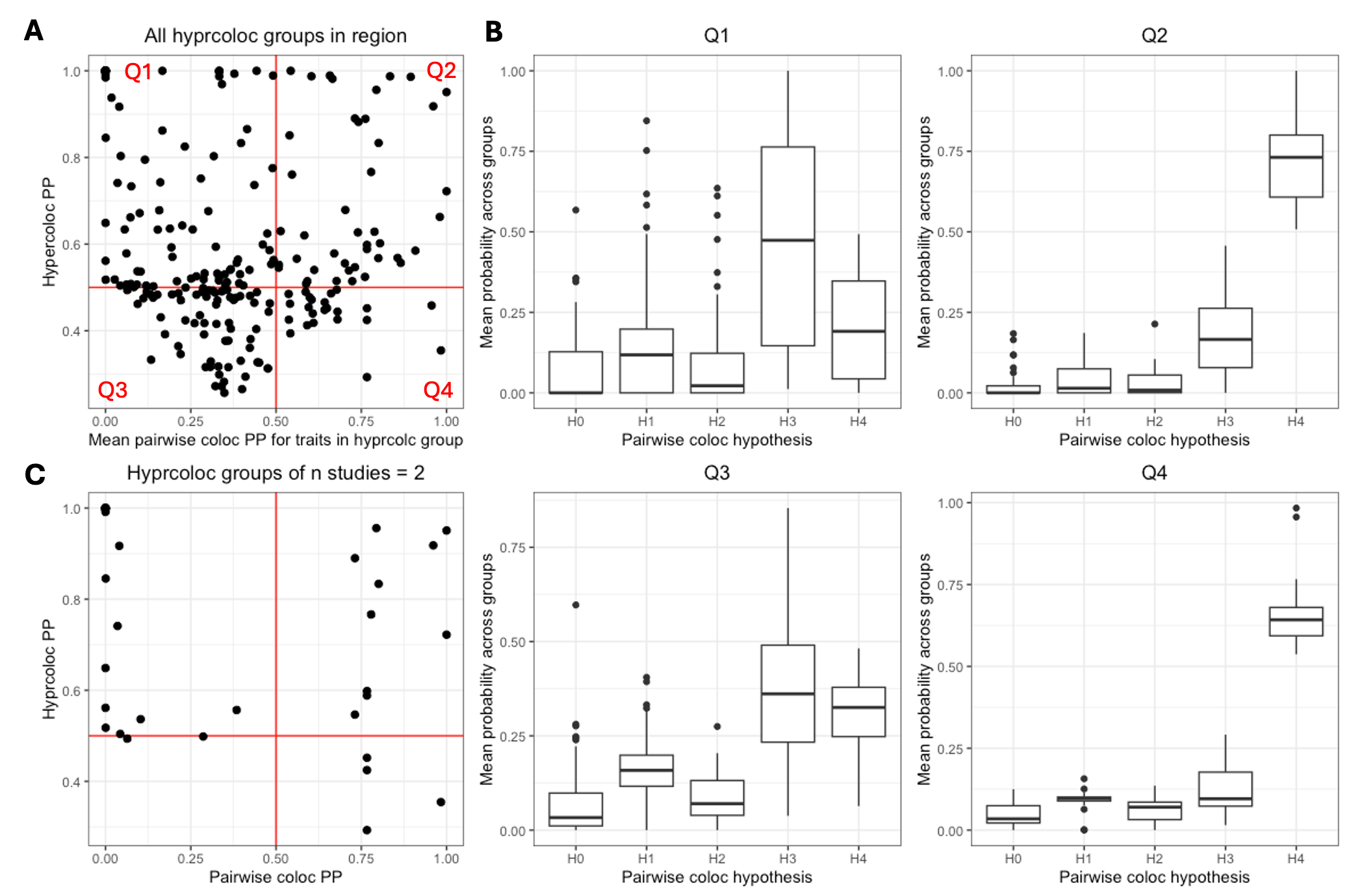

**Figure SN4**: Correlation between HyPrColoc and coloc

Correlation between HyPrColoc posterior probability for groups of colocalizing studies in an LD block on chr 22:35695831-37164130 and the mean coloc H4 posterior probability between each study pair within those groups. The mean coloc posterior probability across all 5 hypotheses tested by coloc for groups of studies in each quadrant is shown in **(B)**. **(C)** An equivalent plot showing only those HyPrColoc groups of n=2 studies. H0 = no association in region, H1 = association at study 1 only, H2 = association at study 2 only, H3 = non-colocalizing associations in region, H4 = colocalizing associations in region.

**Assessing study clustering methods:**

In order to group fine-mapped study regions in each LD block into clusters sharing a colcalising genetic signal, we first generated network graphs using the graph_from_adjacency_matrix function from the igraph package(5). Adjacency matrices (n x n) were constructed from the coloc H_4_ values derived for all pairs of finemapped trait loci (n) in an LD block, setting any H_4_<0.8 to 0 (indicating absence of colocalization), and graphed with edge weighting:

graph_from_adjacency_matrix(H4_adj_mx, mode="undirected", weighted=TRUE, diag=FALSE)

such that an edge linking two traits (nodes) indicated the presence of a high probability colocalizing signal. Upon visualization of resulting network graphs, it was apparent that large clusters of studies could be sparsely linked by individual nodes that joined several more densely connected study subclusters. Assessing the properties of these sparsely connected nodes, they tended to represent studies with higher regional minimum p-values, lower regional SNP coverage and higher average H_0_ coloc results (no genetic signal in the region), indicating the sporadic behavior of colocalization in settings where genetic signals are less robust (Figure SN5-A and B). In order to prune low confidence edges and split apart sparsely connected study subclusters (modules) we trialed the igraph::cluster_edge_betweenness function. This iteratively prunes the graph edge with the highest edge-betweenness score (a measures of the number of shortest paths between nodes that pass through an edge, with high scores resulting from edges that link more densely connected modules(6)), before recalculating edge-betweenness scores and repeating. Edges pruned in order to achieve the maximum modulatory score for the graph (i.e. the maximum number of identified modules) were considered false positive H_4_ results and were deleted along with any resulting unconnected nodes (Figure SN5-C). Linked modules that showed differing study membership but remained connected by unpruned edges were considered separate study clusters (Figure SN5-D). Visualization of region plots for studies clustering within defined modules showed distinct colocalizing signal peaks, indicating the validity of cluster splitting by edge pruning to distinguish independent genetic signals (Figure SN6).

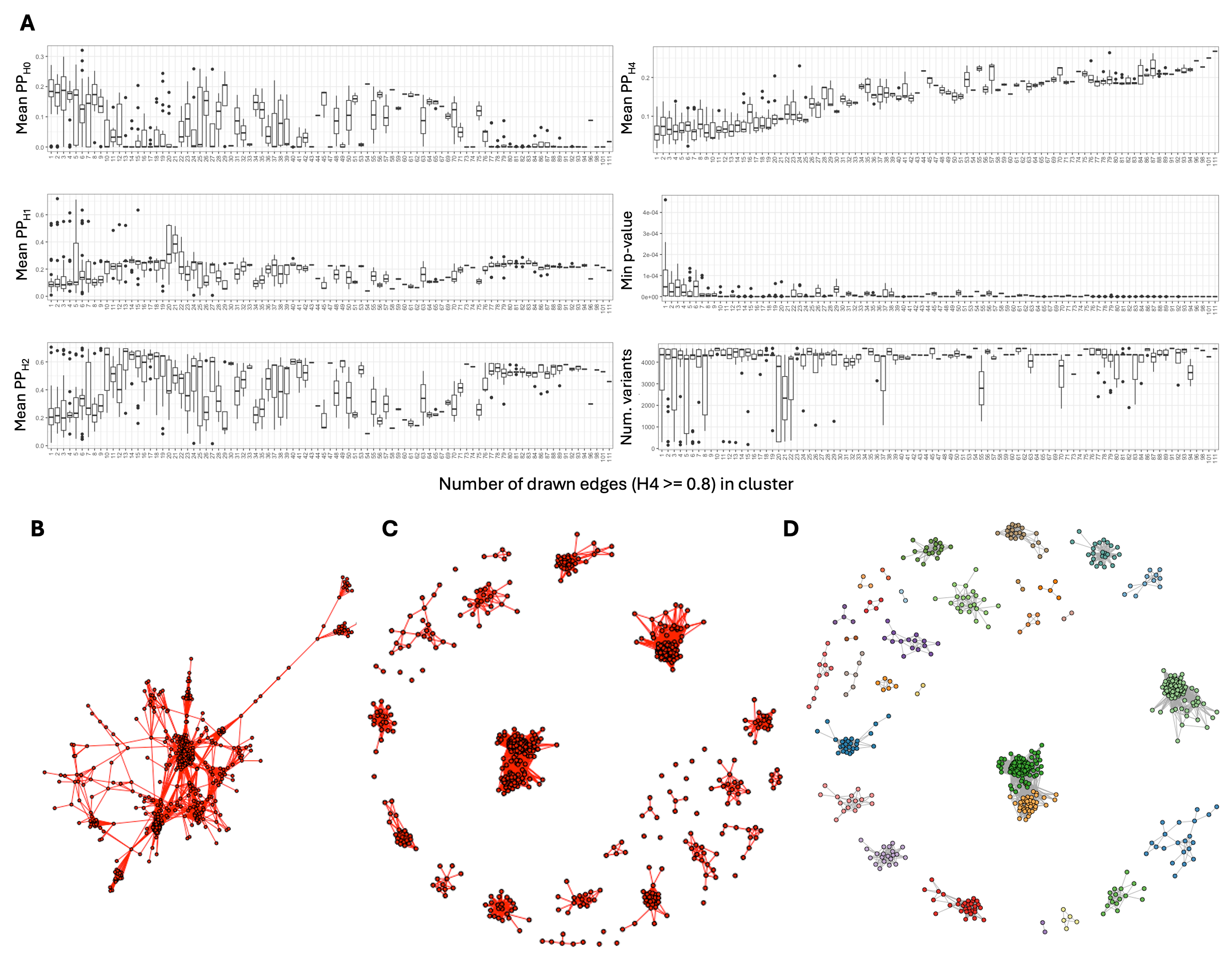

**Figure SN5**: Clustering and Pruning Visualizations

**(A)** The properties of studies (nodes) based on their connectivity to other studies (i.e. number of drawn edges) in the colocalizing network graph shown in B. The mean posterior probability of each tested coloc hypothesis (across all study pairs), the study minimum p-value in the region and the number of variants available for colocalization after fine-mapping are shown. **(B)** An example network graph for a cluster of studies linked by high H_4_ coloc results in the LD block chr22:35695831-37164130. **(C)** The network graph shown in B split into study modules after edge pruning based on edge-betweenness score. **(D)** The same graph with orphaned studies removed and nodes colored by module membership.

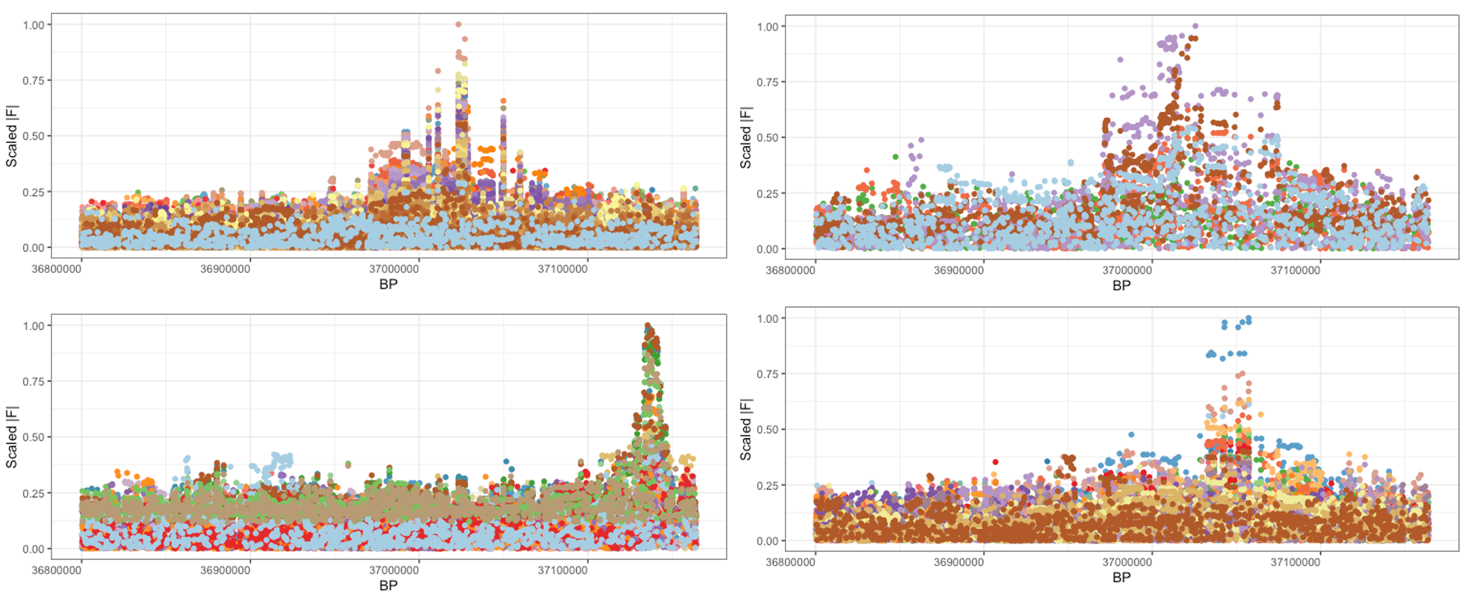

**Figure S6**: Overlayed fine-mapped signals of clustered traits

Example locus plots showing overlayed fine-mapped signals from studies grouped into different modules in the network graph shown in Figure SN5-C/D. Each panel represents a different module, with the studies in that module coloured at random. X-axes show base-pair position on chromosome 22 and y-axes show fine-mapped F statistics scaled as abs(F)/max(abs(F)) across all studies in the module.

Although well performing, we found the computational demand of iteratively recomputing large graph structures and edge-betweenness scores to derive final study cluster assignments to be infeasible for upscaling across the resource. We instead tested the performance of 6 further igraph community detection algorithms (cluster_fast_greedy, cluster_infomap, cluster_label_prop, cluster_leiden_modularity, cluster_louvain and cluster_walktrap) in terms of speed and accuracy in defining independent modules of colocalizing studies within simulated graph structures. Methods were compared in terms of their sensitivity to false negatives (edges absent between nodes in a highly connected module) and false positives (erroneous edges linking separate modules) in correctly assigning nodes to modules. cluster_infomap was most robust to high false positive and false negative rates in the graph structure and ran substantially faster than cluster_edge_betweenness. Based on these parameters, we took this method forward to assess its performance on empirical data.

**Pre-clustering QC, edge weighting and pruning:**

Prior to colocalization, we performed QC on the imputed and fine-mapped results, **see Methods**. If the study region had no further significant associations at our chosen threshold of p<1.5e-4 after variant exclusion the region was dropped. We further screened for spurious colocalizing fine-mapped results from the same original study that showed H_4_**≥**0.8, retaining only the credible set with the smallest minimum p-value.

The cluster_infomap algorithm detects community structure in complex weighted networks by capturing the probability flow of a random walk on that network to decompose it into a series of modules(7). Clustering colocalizing studies into modules by this means enabled putative false positive coloc results spanning modules to be pruned, and putative false negative coloc results linking studies within more densely connected modules to be inferred. We assessed the performance of the algorithm under varying edge weighting strategies by calculating the average H_4_ connectivity score across resulting modules (being the proportion of the maximum number of possible edges in a module that are observed, also known as the module density). We found assigning H_4_ values <0.8 to 0, then scaling the remaining H_4_ values based on the magnitude of the maximum p-value between the two studies in the colocalizing pair, most effectively down-weighted colocalizations between low powered studies to maximize the resulting average H_4_ connectivity of clusters in a region.

The weighting algorithm is defined as such: Given a p-value $p$, we define the edge weight $w(p)$ as:

$$w(p)=\left[ \frac{\text{clamp}(-\log_{10}(p),-\log_{10}(\tau_{1}),-\log_{10}(\tau_{2}))-(-\log_{10}(\tau_{1}))}{-\log_{10}(\tau_{2})-(-\log_{10}(\tau_{1}))} \right]^{\alpha}\cdot(1-w_{\min})+w_{\min}$$

where:

- $\tau_{1}=1.5e^{-4}$ is the upper (lenient) p-value threshold,
- $\tau_{2}=5e^{-8}$ is the lower (genome-wide significance) p-value threshold,
- $\alpha=4$ is the attenuation parameter controlling how rapidly the weight increases between the two thresholds,
- $w_{\min}=0.5$ is the minimum weight assigned to studies at or above $\tau_{1}$,
- $\text{clamp}(x,a,b)=\min(\max(x,a),b)$.

The raw $-\log_{10}(p)$ value is clamped to the interval $\left[ -\log_{10}(\tau_{1}),-\log_{10}(\tau_{2})]=[4,7.3 \right]$, linearly rescaled to $\left[ 0 , 1 \right]$, raised to the power $\alpha$ to attenuate intermediate values, and then mapped to the range $\left[ w_{\min} , 1 \right]$. This yields $w=w_{\min}$ for $p\geq\tau_{1}$ and $w=1$ for $p\leq\tau_{2}$, with a smooth, concave transition in between. The weight is then zeroed out for any pair where $H_{4}<0.8$ the posterior probability threshold, so only colocalized pairs contribute to the adjacency matrix.

Following clustering, we removed all nodes from the cluster that had a <5% amount of connectivity compared to the most connected vertex in the cluster. In rare cases, fine-mapped credible sets from the same trait that didn’t strongly colocalize with each other were pulled into the same module by the clustering algorithm. In these cases, only the credible set with the smallest p-value was retained, though this situation potentially tags clusters of studies with independent fine-mapped signals in very close proximity that are variably called by coloc and should be further investigated.

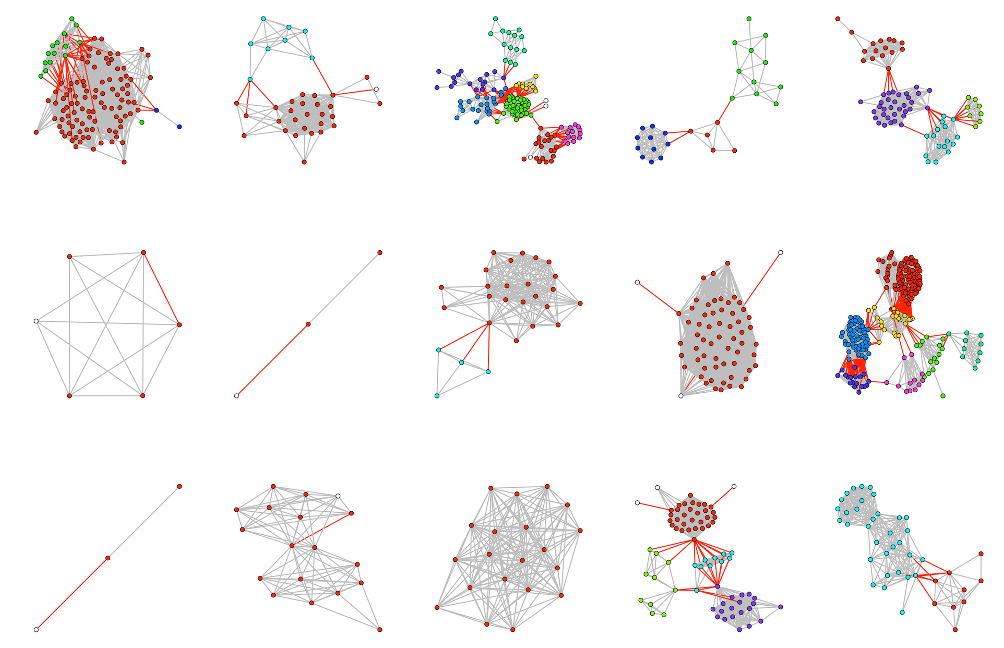

**Figure SN7:** Example network graphs

Example network graphs for studies showing colocalizing signals in the LD block spanning chr1:153358890-156424168, with nodes representing studies and edges representing a colocalizing genetic signal between study pairs at H_4_<=0.8. Independent modules (study clusters) within a graph are distinguished by color and split apart by pruning joining edges (colored red). White nodes indicate studies that are removed post clustering due to low connectivity or being a fine-mapped region from the same origin study as another node in the graph.

A selection of cluster_infomap derived study clusters from the LD block spanning chr1:153358890-156424168 are shown in Figure SN7.

**Validating study clusters:**

We visualized the fine-mapped locus plots for studies clustered by the cluster_infomap algorithm within chr1:153358890-156424168 to 1. assess the legitimacy of colocalizing genetic signal and 2. assess situations in which edge pruning had split apart linked modules for evidence that the underlying colocalizing signals were distinct. Figure SN8 shows example region plots for a densely linked module containing 20 molecular and complex traits from across GTEx, eqtlGen, Brain-eMeta, GoDMC methylation, and GWASCatalog resources, with an annotated shared causal variant at 1:153739486.

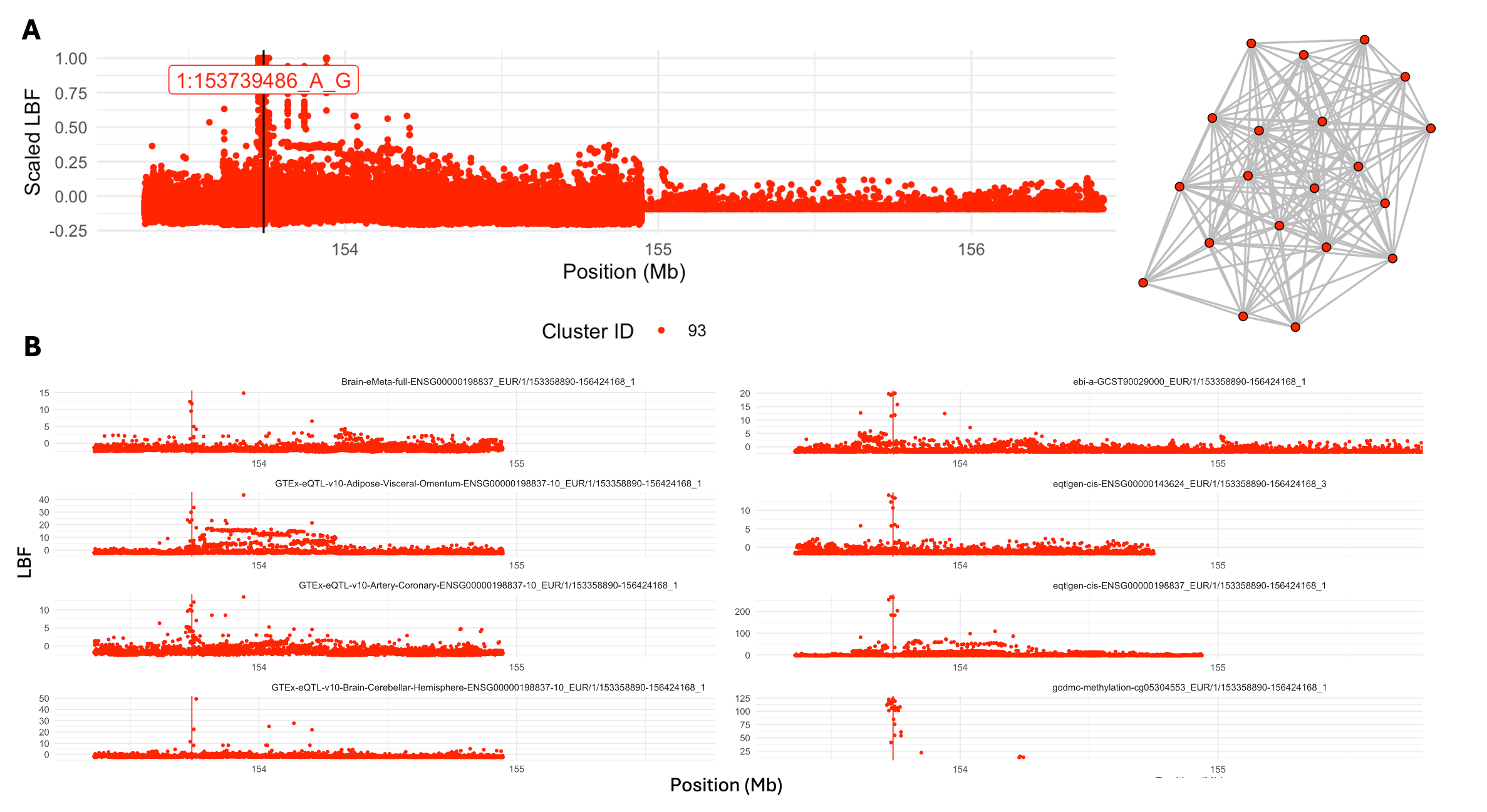

**Figure SN8:** Overlayed locus plot of traits in cluster

**(A)** Overlayed locus plot of all studies in the network graph shown to the right, within the LD block chr1:153358890-156424168. The position of the assigned candidate SNP is shown. X-axis denotes the megabase position of variants on chromosome 1, y-axis indicates the log-bayes factor (LBF) for each variant, scaled within each study as LBF/max(LBF). **(B)** Example locus plots for eight of colocalsing the studies in the cluster; red line indicates the position of the candidate SNP.

Figure SN9 shows example region plots for studies originally linked in a complex graph structure that was split apart by the clustering algorithm into 11 independent modules. Each module shows evidence of colocalization at a differing signal in low to moderately high (r^2^=0.85) LD. (Figure SN10). We could not eliminate the possibility that, in some situations, differences in the performance of fine-mapping and colocalization due to underlying technical differences in the input GWAS data (i.e. low SNP coverage, low power and varying degrees of overlap with the LD reference panel) may result in artificial splitting of study clusters that colocalize. In situations where assigned candidate SNPs for separate study clusters are in strong LD (r^2^=0.99) we report the identity of all potentially overlapping clusters to users.

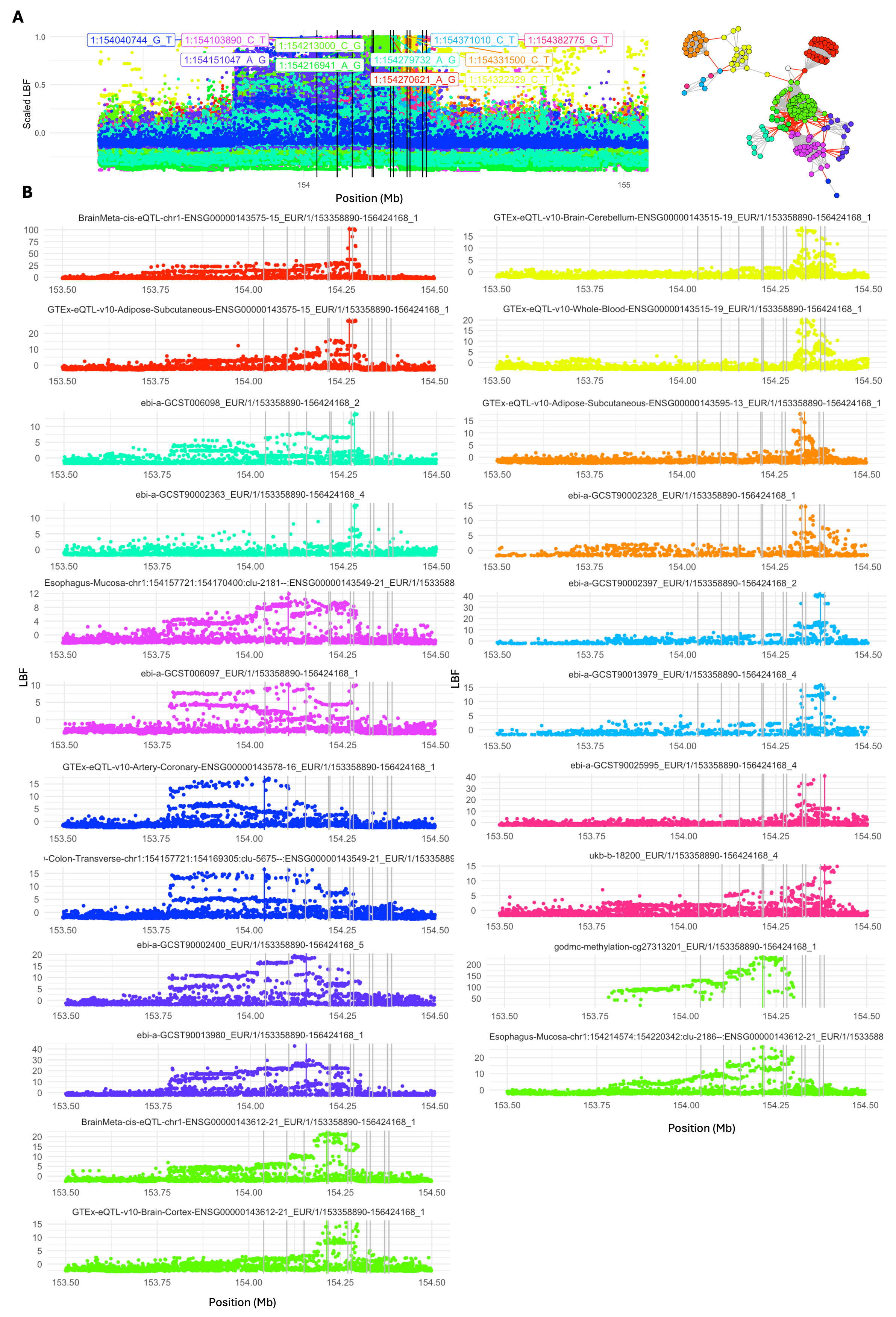

**Figure SN9**: Example of complex cluster pruning

**(A)** Overlayed locus plot of all studies in the network graph shown to the right, within the LD block chr1:153358890-156424168. Studies are colored according to their assigned module, and the position of candidate SNPs are shown by black vertical lines. X-axis denotes the megabase position of variants on chromosome 1, y-axis indicates the log-bayes factor (LBF) for each variant, scaled within each study as LBF/max(LBF). Red edges in the network are pruned to split modules. **(B)** Example locus plots (two from each module) for 22 of the studies in the original network. The position of the assigned colocalizing candidate SNP is indicated by the vertical colored line for each module, with the candidate SNPs of the other modules shown in grey.

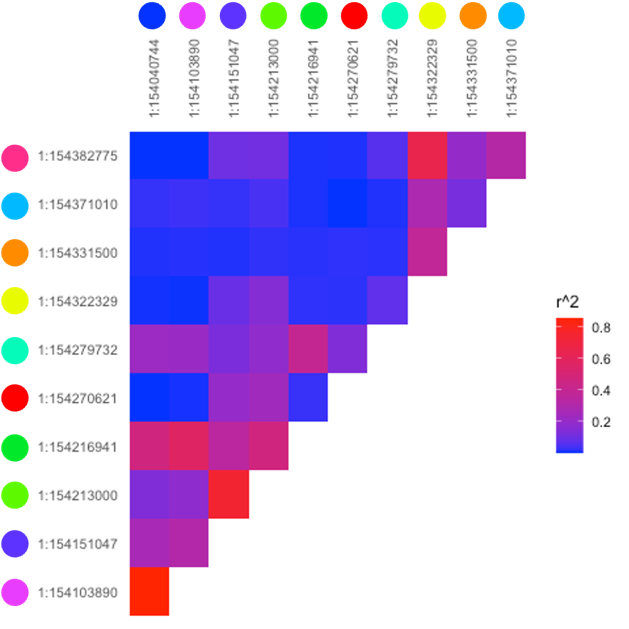

**Figure SN10**: LD between candidate SNPs assigned to the study clusters in Figure SN9

### Supplementary Note 4: Insight into colocalization groups

We have chosen exemplars of different types of colocalization groups that may arise when looking at the GPMap resource.

**Case 1: Low pleiotropy Gene of interest**

We highlight the triggering receptor expressed on myeloid cells 2 (*TREM2*) gene, which is currently under intensive investigation as a putative causal driver and therapeutic target for Alzheimer’s disease*(8)(9)*(see <https://gpmap.opengwas.io/gene.html?id=TREM2>). Our framework provides further empirical support for the prioritized status of *TREM2*; we identify robust colocalization evidence between the gene and Alzheimer's disease across multiple loci, encompassing both common and rare variant architectures.

Notably, our analysis reveals a lack of additional trait associations or secondary gene implications at these loci. This observed lack of pleiotropic effects suggests a highly specific functional role. In a therapeutic context, such a restricted pleiotropic profile is significant, as it indicates a lower probability of inducing off-target deleterious effects or unintended systemic consequences when modulating the gene's function. This high phenotypic specificity, combined with consistent colocalization evidence, reinforces *TREM2* as a high-priority candidate for targeted drug development.

**Case 2: Colocalization Groups in high LD**

Two Colocalization Groups with the same Candidate Variant, but no overlap in pairwise colocalization results.

Here we highlight a single candidate variant (4:112607864 A/G, <https://gpmap.opengwas.io/variant.html?id=6703588>) that was programmatically assigned two distinct colocalization groups. Here, we see that although they share a common candidate variant, none of the studies colocalize with each other.

In Figure SN11 we see the colocalization groups before and after pruning. On the left, we see some low powered loci that link the two groups via significant colocalization results (H_4_ ≥ 0.8), however, on the right, we see that these mediating nodes have been removed, along with other outlier loci that did not colocalize with a significant proportion of the other loci that were colocalizing with each other, and were therefore removed.

This use case necessitates cautious interpretation. The absence of overlapping, high-probability colocalization signals suggest that these associations likely represent distinct genetic architectures driven by divergent biological processes. Although our framework has assigned a shared ‘candidate variant’ for these traits, this may reflect the condensation of independent signals within a region of high linkage disequilibrium rather than a singular pleiotropic mechanism. Consequently, it is incumbent upon the researcher to conduct further analysis to elucidate the precise spatial and functional relationships between these independent colocalization groups.

There are 93,201 total candidate variants among the 97,393 colocalization groups (4192 colocalization groups identified the same candidate variant – but showed inconsistent pairwise colocalizations or had different colocalization patterns). Some resulting candidate variants were also in LD with each other. 8.5% of candidate variants were in high LD (r^2^ > 0.9) and a further 15.2% in moderate LD (r^2^ > 0.8) with another.

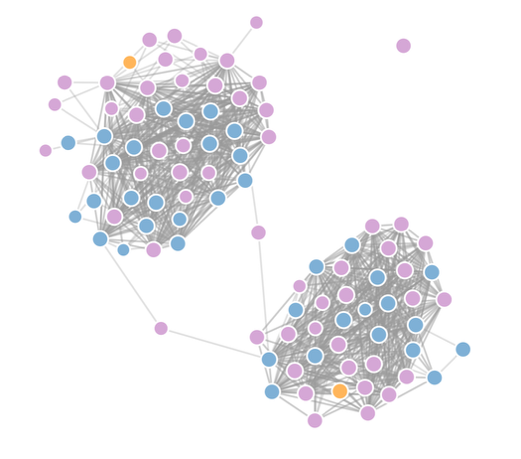
vs.
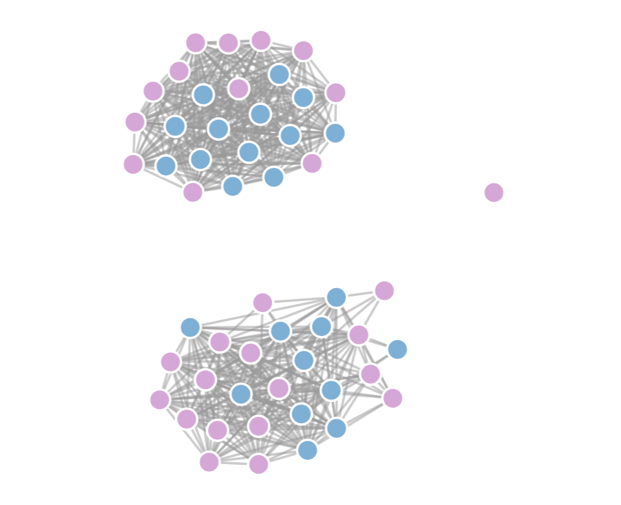

**Figure SN11:** Colocalization Groups with the same candidate variant, before and after pruning.

**Case 3: Colocalization Group with additional low powered traits**

Here we highlight a single candidate variant (6:41066261 G/C, <https://gpmap.opengwas.io/variant.html?id=7829657>) which is assigned to a colocalization group with additional studies that are likely related but were not programmatically included to the colocalization group due to inconsistent pairwise colocalization results. When clustering, we down weight studies which have low power (See Supplementary Note 5), because of this, and the inconsistent nature of pairwise colocalization results for low powered studies, some studies which may be related to the colocalization group which did not make It into the group. In Figure SN12, on the left we see all significant pairwise colocalization results drawn between lower powered trait loci, and the results on the right, where post-pruning we see the same trait loci not included in the final colocalization group.

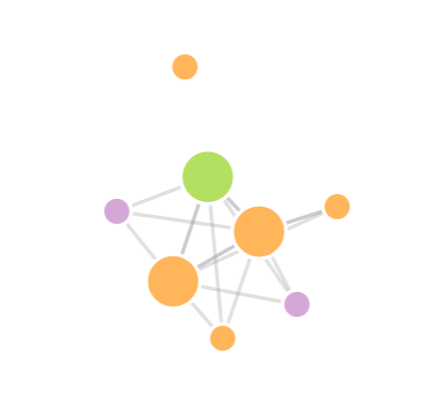
vs.
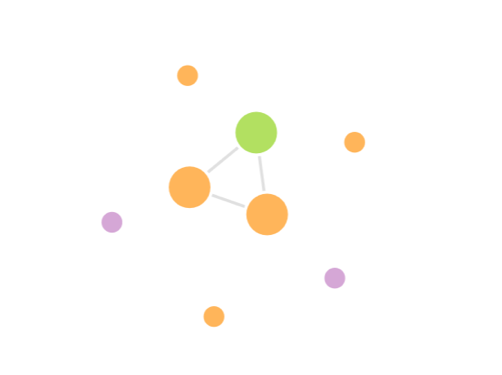

**Figure SN12**: Colocalization Groups with low powered traits

**Case 4: Candidate Variant with many Trait Categories, and few Genes**

This variant (1:167463819 T/C, <https://gpmap.opengwas.io/variant.html?id=468011>) has a colocalization group with one gene tagged by molecular traits, *CD247*, yet is implicated with a series of different traits which have been assigned to different trait categories. This gene encodes the T cell receptor (TCR) CDR3 zeta chain and is essential in TCR signaling to orchestrate immune responses to antigenic stimuli. We see colocalisation of QTLs associated with whole blood methylation and expression of this gene with several immune cell phenotypes related to CDR3 surface expression, and complex immune-related traits including asthma (Physiological measure), eczema (Disease of skin and subcutaneous tissue), white blood cell count (Physiological measure), hypothyroidism (Metabolic disease).

This example illustrates two critical phenomena in the architecture of the GPMap. First, it demonstrates that a variant can exhibit extensive pleiotropy across diverse trait categories without necessarily being detectably pleiotropic across multiple genes. Second, it highlights the inherent complexity in taxonomizing complex traits, which frequently span multiple physiological and clinical domains. To note, the absence of detected gene level cis-pleiotropy at this locus, which has substantial involvement in modifying cell state and transcriptional programmes, could potentially be explained by the high cell type and context specificity of *CD247* expression, and the anticipated trans effects of CD247 signaling on gene expression genome wide. We have not yet derived trans-pleiotropy scores for QTLs, which would potentially provide far greater insight into the consequences of variation in cell surface receptor signaling.

The classification of Asthma provides a representative case study of trait category ambiguity. While biologically it could be considered an immune-mediated or respiratory disease, categories which are not currently represented in our high-level taxonomy, it could also be viewed as an environmental measure due to its significant exogenous component. However, in many large-scale repositories, such as the UK Biobank, asthma is frequently categorized under Physiological Measures as it is often defined by quantitative spirometry (e.g., FEV1/FVC ratios) or peak flow assessments. Consequently, we have aligned our classification with this convention.

Furthermore, certain traits remain vaguely defined, potentially encompassing multiple distinct diseases and classifications. For instance, the trait “*Trait: Blood clot, DVT, bronchitis, emphysema, asthma, rhinitis, eczema, allergy diagnosed by doctor: Hayfever, allergic rhinitis or eczema*” (Disease Of Skin And Subcutaneous Tissue) spans several organ systems. In cases where the definitions were too broad to permit meaningful biological grouping, such as “*Trait: Blood clot, DVT, bronchitis, emphysema, asthma, rhinitis, eczema, allergy diagnosed by doctor: None of the above*” (N/A), we elected to leave the trait unclassified.

Despite the inherent difficulties in categorizing disease aetiology, these designations provide a valuable high-level heuristic for assessing the pleiotropic breadth of a variant. By simplifying complex phenotypic landscapes, this classification allows researchers to prioritize loci for further investigation into their specific causal mechanisms with greater clarity.

**Case 5: Candidate Variant implicated with many Genes, but few trait categories**

This example variant (1:156275454 T/C, <https://gpmap.opengwas.io/variant.html?id=426023>) illustrates the opposite of case 3, whereby a colocalization group exhibits high gene pleiotropy, but little trait category pleiotropy. This might interest a researcher who is curious about the interactions of a cluster of genes (in this case *PAQR6, SMG5, GLMP, CCT3, SEMA4A, BGLAP, TMEM79, PMF1, SLC25A44*). They can investigate the region in which all these genes are proximal and are putatively controlled by similar regulatory variants, yet do not have a wide range of pleiotropic effects and are only linked to some physiological blood measures (Albumin, Triglycerides, Basophil).

### Supplementary Note 5: Differential representation of complex traits amongst BMI colocalization groups stratified by tissue annotation

The results of principal component analysis indicated evidence for differences in the complex traits represented in BMI colocalization groups annotated by tissue eQTL, highlighting axes of separation between whole blood, brain and subcutaneous adipose tissue (**Figure 5c**). The candidate variants for BMI annotated by brain- and subcutaneous adipose tissue were next incorporated into a multivariable MR framework as separate exposures (brain-BMI and adipose-BMI respectively). Importantly, the incorporation of stratified sets of IVs into downstream analyses such as tissue-partitioned multivariable MR is dependent on our confidence that the stratification procedure has identified distinct effects underlying the measured exposure. We investigated this further, aiming to demonstrate how candidate variants stratified by tissue-annotation may help identify sets of IVs enriched for differential biological processes.

To achieve this, we applied hierarchical clustering to z-scores calculated from within-tissue proportions of variant–trait associations, considering all complex traits tagged by the set of BMI colocalization groups. Patterns of trait enrichment between tissues were visualized by generating a heatmap (**Figure SN13**). The results of this analysis identified sets of traits which are more enriched within BMI colocalization groups annotated by a given tissue compared to other tissues. For whole blood, this yielded a broad set of traits associated with BMI capturing a number of systemic processes including circulating measures of lipids, inflammation, hepatic pathways cardiovascular disease and longevity. Given the highly pleiotropic signal of candidate variants annotated by whole blood, these were not considered as a distinct effect for inclusion in downstream MR analyses. The traits enriched within brain-annotated colocalization groups depict both centrally regulated processes, such as behavior (e.g. smoking status, alcohol intake), mood (e.g. depressed affect, feeling miserable) and heart rate; and an adverse cardiometabolic profile indicated by visceral adipose tissue volume and cardiovascular disease outcomes. In contrast, the traits enriched within subcutaneous adipose-annotated colocalization groups largely comprised measures of lipoprotein composition and lipid metabolism. The full set of traits which provided evidence for enrichment (indicated by a Z>2) for each tissue is provided in **Supplementary Table 8**).

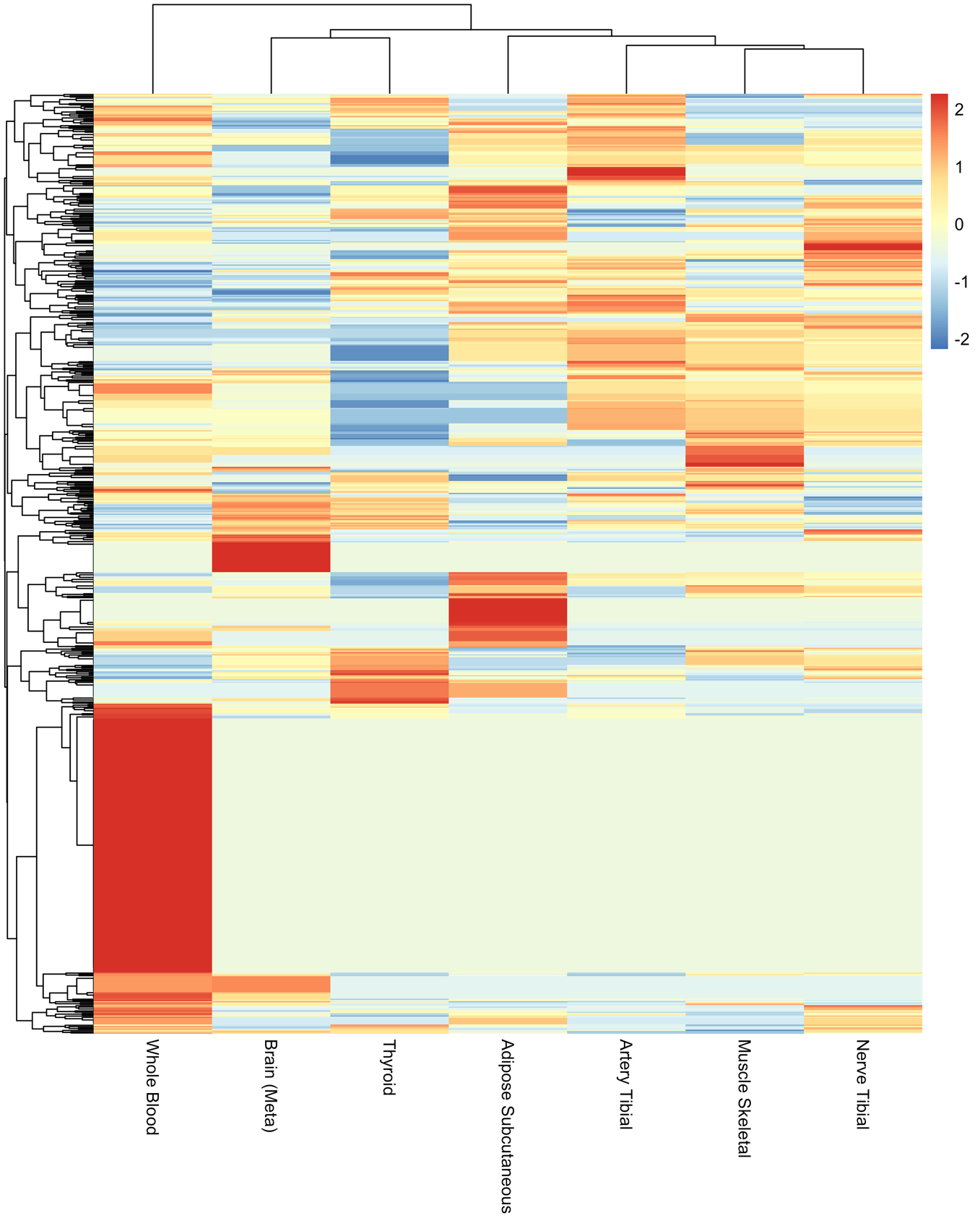

**Figure SN13:** Heatmap illustrating differential enrichment of traits between tissue-annotated candidate variants for BMI

The heatmap represents the relative enrichment of complex traits within BMI colocalization groups per tissue after normalizing for the total number of complex trait associations per tissue-annotated colocalization group. Values are shown as z-scores across tissues, such that higher values denote greater relative representation, with z-scores > 2 highlighting enriched signals.

### Supplementary Note 6: Limitations of input and statistical methods used in the GPMap pipeline

Limitations due to Quality of GWAS studies / Quality Control / Legitimacy of Results

The GPMap resource can facilitate the selection of IVs for exposures of interest in MR analyses by selecting candidate SNPs representing CG groups. We recommend that analysts are aware of several sources of bias in IV selection which should be considered when refining IV selection. For example, analyses should consider excluding highly pleiotropic candidate variants, or variants sharing CG with certain GWAS traits which may act as an indicator of spurious association due to population stratification and selection bias.

**Limitations due to Linkage Disequilibrium**

The impact of between population LD structure on colocalization is a challenge when aggregating summary statistics from different studies and populations. While coloc(2) doesn't explicitly require an LD matrix as input (it infers it implicitly from the pattern of association across SNPs), accurate LD patterns are critical for reliable results. Further, imputation and fine-mapping explicitly require an LD matrix, for which we have used a single LD matrix from the UK Biobank. Ideally, each source of summary statistics would be accompanied with an LD matrix in which imputation and fine-mapping could be performed. However, that is currently not available.

There are current challenges in fine-mapping highly dense genomic regions, where LD structures are complex (e.g., the HLA/MHC locus). In such regions, fine-mapping does not converge at a higher rate. Further work could be done to refine fine-mapping methodologies to improve causal variant resolution.

**Limitations of Colocalization Methods and Clustering**

The robustness of colocalization analysis is intrinsically dependent on the quality of the input summary statistics derived from the source studies. Critically, any systemic biases present in these original data, such as uncontrolled confounding or technical batch effects, are directly propagated into the estimated colocalization probabilities. Further, limitations of both power and genome coverage influence the number of colocalizations found, however we have mitigated these limitations by imputing all summary statistics in the GPMap. Many of the tissue specific QTL data have a limited sample size, and traits with primary biology in an inaccessible tissue will show less colocalizations than those with a significant circulatory component to their biology.

While colocalization methods often assume a single causal variant, we attempted to mitigate this constraint by incorporating fine-mapped results prior to colocalization testing. However, this introduces a dependency: if the fine-mapping procedure produces spurious credible sets, the resulting colocalization probabilities will be similarly compromised.

A key limitation observed was the lack of consistency in pairwise colocalization results. Specifically, the transitive property, where the colocalization of A with B and A with C implies the colocalization of B with C, was often inconsistent. This inconsistency is primarily driven by how preceding analytical limitations or limited statistical power manifest as heterogeneity in pairwise colocalization analyses. This phenomenon introduces challenges when clustering results into colocalization groups, leading to both false positive and false negative inferences. We addressed this issue by developing a specific adjustment methodology (**Supplementary Note 5**).

### Supplementary Note 7: Pipeline Architecture

The GP Map is comprised of 3 distinct software repositories, *genotype-phenotype-map*, which is comprised of the code to process the GWAS data, which includes all steps in Figure 1. The code in genotype-phenotype-map creates the databases used in the API and is also repackaged and deployed on a web server, which allows for users to upload their own GWAS for processing. *genotype-phenotype-api* comprises both the API and the website, which presents the results of the GP Map. *gpmapr* is an R package that allows for programmatic access to the data and allows for future statistical methods to be built on top of the data.

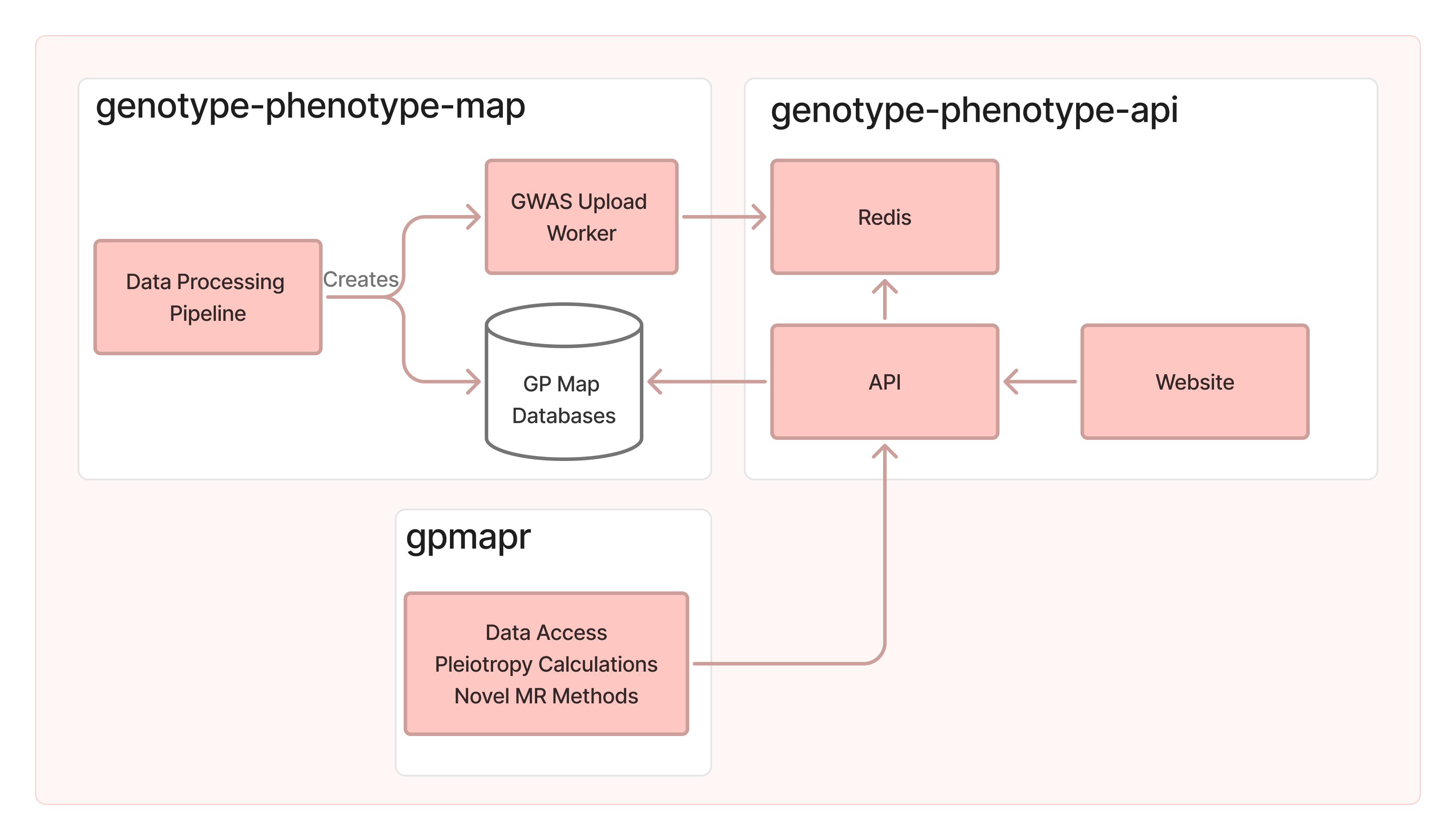

**Figure SN14:** Genotype-Phenotype Map Architecture Diagram

Each white box represents a different code repository, genotype-penotype-map is the data processing pipeline, which also doubles as the GWAS Upload functionality. genotype-phenotype-api encapsulates the API, website, and server orchestration. gpmapr is the R package to access the GP Map data, and to provide a future layer of statistical methods that can use the underlying data.

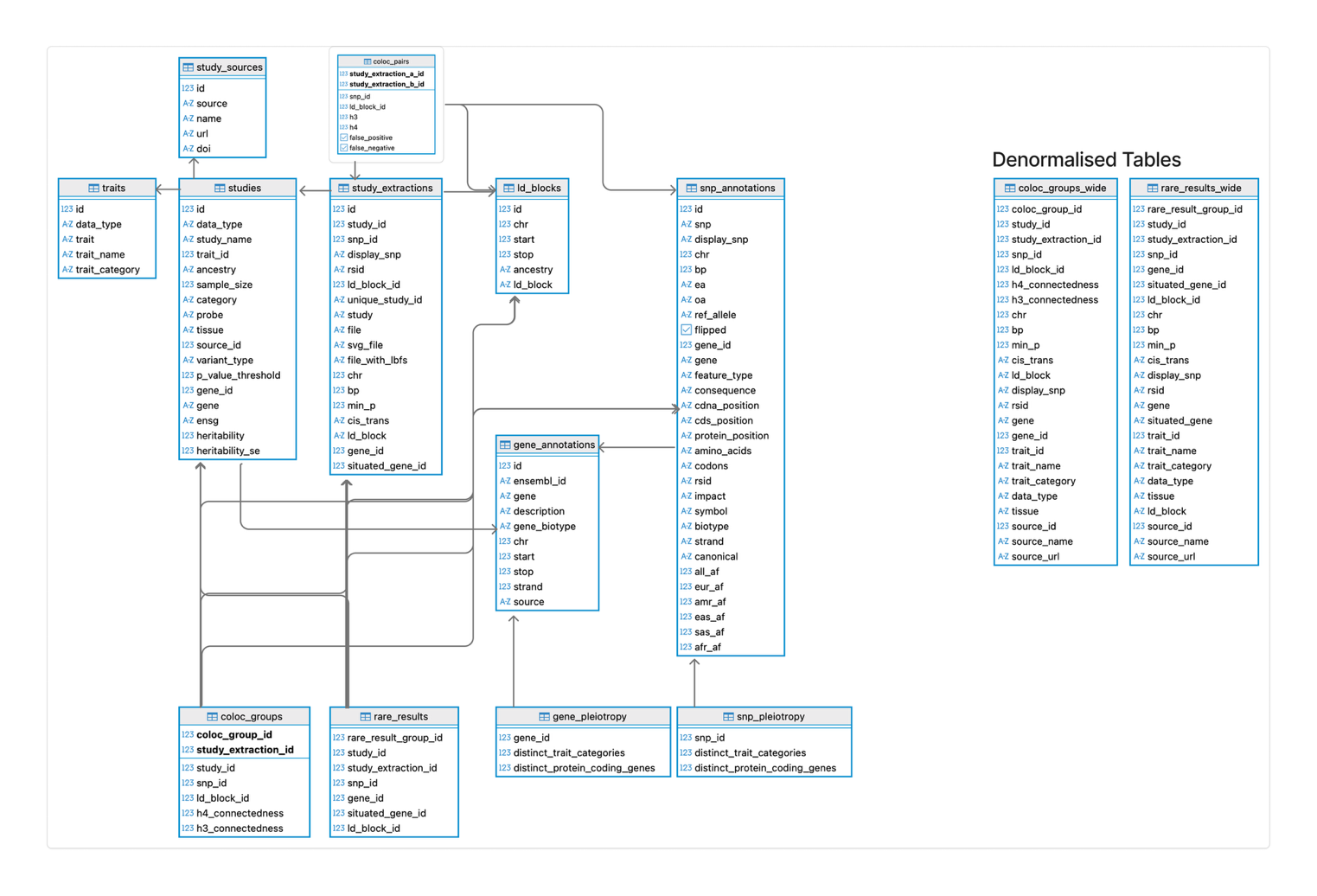

**Figure SN15**: Genotype-Phenotype Map Database Schema

Each arrow represents a relationship between tables, which show how each table and concept is related to another.

### Supplementary Note 8: Imputation of GWAS summary statistics

Effective imputation methods exist for GWAS summary statistics against LD reference panels (ssimp(10) and RAISS(11)) however they are computational demanding due to the requirement of matrix inversion. We introduced an alternative approximate method for imputation of missing summary statistics against an LD reference panel that uses pre-computed eigenvectors of LD matrices for pre-defined LD regions, thus substantially increasing the speed of the imputation process and making the pipeline more computationally tractable.

For a vector of $\boldsymbol{b}$ true causal effects for $M$ variants, the expected marginal effect estimates are

$$\boldsymbol{\beta=}\boldsymbol{D}^{\boldsymbol{-}\frac{\boldsymbol{1}}{\boldsymbol{2}}}\boldsymbol{\rho}\boldsymbol{D}^{\frac{\boldsymbol{1}}{\boldsymbol{2}}}\boldsymbol{b}$$

Where $\boldsymbol{\rho}$ is an MxM LD correlation matrix and the diagonal matrix $\boldsymbol{D}$ contains the variances of each variant based on allele frequency $p_{j}$, $D_{j,j}=2p_{j}(1-p_{j})$. Variances of variant effect estimates are obtained from $cov\left( \hat{\boldsymbol{\beta}} \right)\boldsymbol{=S\rho S}$ where $\boldsymbol{S}$ is a diagnonal matrix with $S_{j,j}=\sigma_{j}\approx\sqrt{var(y)/2p_{j}\left( 1-p_{j} \right)N}$, and $\boldsymbol{z=}\hat{\boldsymbol{\beta}}\boldsymbol{/}\boldsymbol{\sigma}_{\boldsymbol{j}}$. If some values of $\boldsymbol{z}$ are missing ($\boldsymbol{z}_{\boldsymbol{i}}$) we wish to project the relationship between observed $\boldsymbol{z}$ values ($\boldsymbol{z}_{\boldsymbol{t}}$) and $\boldsymbol{\rho}$ onto the missing values. To approximate this we perform principal component analysis of the LD matrix for the region (based on LDetect boundaries) to obtain a matrix of Eigenvectors $\boldsymbol{X}$, and retaining the set of vectors that explain 90% of the variance fit the linear model $\boldsymbol{z}_{\boldsymbol{t}}\boldsymbol{=a+}\boldsymbol{X}_{\boldsymbol{t}}\boldsymbol{\gamma+e}$ using elastic net regularization provided by the R package glmnet(12), with $\alpha=0.5$ and $\lambda$ parameter estimated using 10-fold cross validation. Finally, we estimate the missing values with ${\hat{\boldsymbol{z}}}_{\boldsymbol{i}}\boldsymbol{=}\boldsymbol{X}_{\boldsymbol{i}} \hat{\boldsymbol{\gamma}}$.

Subsequent steps are performed to improve interpretability and reliability.

**Scaling**

Scaling the results of the z-score and standard error estimations uses the following steps:

Outliers are calculated for the ratios of known / predicted where any value outside 3 standard deviations from the median is ignored. From there, a polynomial regression of the known values against the predicted values is taken, which allows us to capture potential non-linearity. We then take the coefficients of the polynomial regression and adjust the predicted values.

**Filtering**

We only use the imputed results if the correlation of effect estimates between the imputed and known values is above 0.7.

From the imputed results, we wanted to be sure that no overinflated p-values could be used for the subsequent steps in the pipeline. For imputed variants with p-values < 1.5e-4, if known variants that are correlated to the imputed variant ($R^{2}>0.6$) are more significant than the non-imputed rows by 10 we removed that variant.

**Data and Code Accessibility:**

The code is available in the GitHub repository <https://github.com/MRCIEU/genotype-phenotype-map>. The PCs of the LD matrix is available in the publicly available Google Cloud bucket <https://console.cloud.google.com/storage/browser/genotype-phenotype-map>.
